# Meta-analysis of glucose metabolism across Alzheimer’s, Parkinson’s and ALS reveals distinct patterns of cortical hypometabolism versus subcortical hypermetabolism with disease-specific spatial signatures

**DOI:** 10.64898/2026.04.07.26350339

**Authors:** Adam C. Raikes, Melissa Garza, Coco Tirambulo, Angela N. Murrell, Roberta Diaz Brinton

## Abstract

The purpose of this meta-analysis was to determine the common signature of dysregulated brain glucose metabolism in late-onset age-associated neurodegenerative diseases (A^2^NDs), including Alzheimer disease, Parkinson disease, and amyotrophic lateral sclerosis. FDG-PET studies published through July 2025 were identified from MEDLINE, Embase, PsycINFO, Scopus, and Cochrane databases. Studies comparing adults with late-onset neurodegenerative disease to healthy controls and reporting whole-brain coordinate findings were included. Activation likelihood estimation meta-analyses were conducted using extracted coordinates of hypo- and hypermetabolism. A total of 130 studies were included, comprising 5,298 individuals with A^2^ND and 3,499 controls. Meta-analyses revealed dysregulated glucose metabolism as a unifying feature across A^2^NDs and included both hypo- and hypermetabolic patterns. However, the spatial distribution of metabolic abnormalities differed across diseases, with each disease demonstrating distinct neuroanatomical patterns associated with disease-relevant functional systems. While hypometabolism primarily in the cortex remains a primary focus in neurodegenerative disease research due to functional relevance, hypermetabolism was also consistently identified across all three diseases, predominantly in subcortical, limbic, and cerebellar structures. This dissociation, with hypermetabolic regions localized to structures with known functional connectivity to affected cortical networks, suggests compensatory upregulation as a plausible interpretation, though alternative mechanisms, including neuroinflammation, cannot be excluded.

---

Collectively, the findings indicate that altered brain bioenergetics are shared across neurodegenerative diseases with unique disease-specific metabolic hypo- and hypermetabolic signatures. These findings highlight the potential importance of addressing bidirectional metabolic dysregulation alongside disease-specific pathological drivers in future prevention and treatment strategies.

## Background

The brain is the most energetically demanding organ in the body, comprising just 2% of body weight while consuming approximately 20% of glucose[1,2]. This high metabolic demand makes it particularly vulnerable to even modest reductions in glucose availability and ATP generation. Such shifts are particularly evident in Alzheimer’s disease, where diminished glucose availability[3,4] induces a shift from glucose metabolism toward fatty acid metabolism[3–13]. Concomitant disruption of mitochondrial bioenergetics[3,8,14–16] and an accompanying neuroinflammatory cascade[7,17–19] culminate in late- onset age associated neurodegenerative diseases[2], including Alzheimer’s disease (AD), Parkinson’s disease (PD), Amyotrophic Lateral Sclerosis (ALS) (hereafter abbreviated A^2^ND for brevity), and Multiple Sclerosis (MS). Deficits in these bioenergetic processes are well-documented hallmarks, emerging before most functional impairments become apparent[20–22]. This indicates that bioenergetic dysfunction is a common feature of A^2^ND with recent work indicating that metabolic dysregulation may in fact be a underlying mechanism for pathogenesis in these diseases[2,23].

Recent disease-specific systematic reviews and meta-analyses of brain glucose uptake using ^18^F-fluorodeoxyglucose positron emission tomography (FDG-PET) consistently identify hypometabolic patterns in AD[24,25], PD[26], and ALS[27]. Hypometabolism is the most commonly reported disease-related metabolic outcome and is often localized to the posterior cingulate, precuneus, angular gyri, motor cortex, frontal cortex, and subcortically in the basal ganglia and thalamus[24–27].

Hypermetabolism, while less commonly reported across these diseases, has been observed in subcortical regions, the cerebellum, and the brainstem[26,27]. These meta-analyses have focused on individual diseases and often focus on the hypometabolic outcomes despite considerable heterogeneity in the patterns of both hypo- and hypermetabolic findings that often depend on the type of analysis at the individual study level (e.g., region-of-interest, voxel-wise, and network-based approaches)[28–36].

On the premise that bioenergetic dysfunction is a common neurodegenerative feature and may be a common disease-driving mechanism[23], it follows that there may be a transdiagnostic spatial pattern to glucose dysregulation in the brain that then further differentiates into disease-specific manifestations. To date, there are no whole-brain FDG-PET meta-analyses that have examined transdiagnostic A^2^ND FDG-PET outcomes in a unified manner to determine both shared and disease-specific spatial patterns of metabolic disruption and their potential behavioral correlates. To address this gap, the following analyses were conducted: 1) spatial localization of A^2^ND transdiagnostic metabolic dysregulation across FDG-PET studies comparing individuals with A^2^NDs to healthy individuals; 2) determination of shared patterns of statistically significant metabolic dysregulation versus disease-specific patterns; and 3) association of functional correlates of brain regions with FDG-PET patterns of glucose uptake differences. These analyses were followed by a coordinate-based meta-analysis to identify spatial convergence of glucose uptake patterns across a wide and diverse literature base of FDG-PET studies. Activation likelihood estimation (ALE) models were applied to case-control studies to identify both transdiagnostic and disease specific bioenergetic patterns. ALE conjunction and subtraction analyses were conducted across individual diseases in a pairwise manner to identify shared and unique spatial patterns. Finally, generalized correspondence latent Dirichlet allocation (GC-LDA) meta-analytic decoding to describe brain networks and functions likely to be affected by neurodegeneration-related glucose dysregulation was conducted.

## Methods

A protocol was developed a priori and registered with PROSPERO (PROSPERO 2023 CRD42023469898).

### Inclusion and Exclusion criteria

*A priori* inclusion criteria specified peer-reviewed human neuroimaging studies 1) of adults with an average age 50 years or older to reflect the late-onset, rather than early onset, A^2^ND profile; 2) report of cerebral glucose metabolism; 3) in individuals with diagnosed or clinically assumed late-onset forms of Alzheimer’s disease, Parkinson’s disease, amyotrophic lateral sclerosis, or multiple sclerosis. Eligible studies were case–control designs using FDG-PET to quantify disease-related differences in cerebral glucose metabolism comparing individuals with an A^2^ND diagnosis to neurologically healthy control participants. Cross-sectional studies were included and longitudinal studies were considered as well providing that PET outcomes were collected after diagnosis (i.e. changes over time within individuals with pre-existing A^2^ND diagnoses at the time of baseline imaging). Studies were required to report cluster-level results in a standardized stereotactic space (Montreal Neurological Institute or Talairach), including peak coordinates, cluster extent, and an associated statistical measure (e.g., *t*-, *F*-, or *p*-values). Excluded reports included clinical trial descriptions without data, systematic reviews and meta-analyses, preclinical or non-human studies, investigations lacking a non-disease control group, studies not using FDG-PET to assess case–control metabolic differences, studies conducted on genetically determined or early onset disease forms, longitudinal studies where only pre-diagnosis imaging was conducted (i.e., studies where individuals with mild cognitive impairment were imaged and then followed-up with after 5 years to classify as converters or non-converters were excluded), and reports that did not provide sufficient information to estimate coordinate-based activation likelihood estimation maps.

### Database search

A systematic literature review was developed and conducted by a librarian (ANM) in November 2023. Databases searched included MEDLINE via Ovid (MEDLINE® ALL, 1946 to 2023), Elsevier Embase.com (including Embase, MEDLINE, Embase Classic, 1947 to 2022), APA PsycINFO (1800-2023), Elsevier Scopus (1788 to 2022), and Wiley Cochrane Library CENTRAL Register of Controlled Trials (Issue 11 of 12, November 2023). Results were limited to English language only, due to the limitations of the team. References lists of reviews were screened for eligible studies. The search terms included controlled vocabulary and keywords for four neurodegenerative diseases Parkinson Disease, Amyotrophic Lateral Sclerosis, Alzheimer Disease, and Multiple Sclerosis, Fluorodeoxyglucose F18, glucose, and positron-emission tomography (PET). The search was developed in MEDLINE and translated to other databases. See Supplement A for the complete search strategies for all five databases. An update was conducted in July 2025.

Records were retrieved and exported to EndNote 21 and deduplicated. Initial screening was conducted using the DistillerAI tool. Three researchers (ACR, MG, CT) double-screened a total of 25% of the records by title and abstract and then trained the DistillerAI tool to screen the remainder. The three researchers then acted as a second screener on the remaining 75% of the records and conflicts between DistillerAI and the researchers were discussed amongst the team. Full texts were then retrieved for potentially relevant articles and were double-screened by three researchers using the specified inclusion/exclusion criteria.

Data from full texts meeting inclusion criteria were extracted in *neurosynth compose* (https://compose.neurosynth.org)[37]. Extracted data included cluster peak and subpeak coordinates, clusterwise t- or Z-values, and annotations indicating the disease of interest, whether the outcome was for hyper- (disease group > control) or hypometabolism (disease group < control), and sample sizes for each participant group in an analysis. No studies in patients with MS fulfilled the inclusion criteria. We retained reference to MS in the description of the inclusion criteria and database search for consistency with our preregistered protocol. However, no analyses were conducted and no interpretations related to MS are provided hereafter. Study-level summaries, including mean ages, sample sizes, and number of hypo- and hypermetabolic foci are reported in Supplemental Table 2.

### Statistical analyses

#### Activation Likelihood Estimation

Patterns of hypo- and hypermetabolism were examined separately across the three included diseases collectively and within each disease individually. For each analysis, an activation likelihood estimation (ALE) meta-analysis[38–40] was performed with NiMARE 0.9.0 (RRID:SCR_017398)[41] using an ALE kernel to generate study-wise modeled activation maps from coordinates. In this kernel method, each coordinate is convolved with a Gaussian kernel with full-width at half max values determined on a study-wise basis using the study sample sizes according to the formulae provided in Eickhoff (2012)[38]. When required, NiMARE converted Talairach coordinates to MNI space. For voxels with overlapping kernels, the maximum value was retained. ALE values were converted to p-values using an approximate null distribution[38]. Family-wise error rate correction was performed using a Monte Carlo procedure. In this procedure, null datasets are generated in which dataset coordinates are substituted with coordinates randomly drawn from the whole-brain meta-analysis mask, and maximum values are retained. This procedure was repeated 10000 times to build null distributions of summary statistics, cluster sizes, and cluster masses. Statistically significant clusters were defined using edge-wise connectivity with a voxel-level threshold of *p* < 0.001 from the uncorrected null distribution and family-wise error corrected cluster-forming *p* < 0.05 threshold[42,43].

#### Disease-specific signatures

To identify shared signatures of metabolic disruption, we conducted pairwise conjunction analyses using NiMARE. Analyses were performed separately for hypermetabolic and hypometabolic contrasts using the disease-specific family-wise error corrected Z-valued cluster maps. At each voxel, the conjunction was defined by retaining the minimum statistically significant Z-value across maps[44].

Conjunction analyses were first performed across all three diseases simultaneously, followed by all pairwise combinations. For all conjunction analyses, statistically significant voxels were identified as those having joint significance across all maps being considered (e.g., all three diseases when considering all three, both diseases in pairwise analyses). The resulting conjunction value was defined as the smaller of the two group-specific Z-values.

To identify unique signatures of metabolic disruption between diseases, pairwise ALE subtraction analyses[45] were conducted using the approach implemented in NiMARE. Pre-specified analysis was a subtraction comparison between the overall map (A^2^ND) and individual diseases. However, NiMARE’s approach generates a permutation-based null p-value distribution by randomly shuffling the labels for the groups. Having diseases represented in both the A^2^ND dataset and individual disease dataset would violate the exchangeability assumption, and thus pairwise disease subtractions fit separately for hypo- and hypermetabolism was conducted. In this approach, ALE-difference scores were calculated between each pair of disease datasets, for all voxels in the whole-brain meta-analysis mask. Voxel-wise null distributions of ALE-difference scores were generated via a randomized group assignment procedure, in which the studies in the two datasets were randomly reassigned and ALE-difference scores were calculated for the randomized datasets. This randomization procedure was repeated 10000 times to build the null distributions. The significance of the original ALE-difference scores was assessed using a two-sided statistical test. The null distributions were assumed to be asymmetric, as ALE-difference scores will be skewed based on the sample sizes of the two datasets.

There is no currently well-established multiple comparisons correction approach for ALE subtraction analyses[46]. Following recommendations in Eickhoff et. al (2011) and the approach implemented in other work[47] clusters were defined using an uncorrected voxel-level threshold of *p* < 0.001combined with a minimum cluster extent of at least 25 connected voxels (200mm^3^). The voxel-level threshold of *p* < 0.001 is consistent with the threshold applied to the primary ALE analyses prior to cluster-level family-wise error correction, while the cluster extent requirement reduces the likelihood of isolated false positive voxels surviving. The permutation-based null distribution provides a non-parametric basis for voxel-level significance testing, but does not yield formal cluster-level family-wise error control.

Therefore, subtraction results should be interpreted with more caution than the primary disease-specific and conjunction ALE findings, which are fully FWE-corrected.

#### Meta-analytic Decoding

To characterize plausible functional and behavioral correlates of the identified patterns of hyper- and hypometabolism, we conducted generalized correspondence latent Dirichlet allocation (GC-LDA) topic modeling[48] adapted from the surface-based framework described by Peraza et al. (2024)[49]. This approach identifies statistical spatial correspondences between metabolic abnormality maps and functional terminology from the broader neuroimaging literature. It does not directly measure function or impairment in the disease populations studied. We used the Neurosynth version 7 data release (http://www.github.com/neurosynth/neurosynth-data), which includes activation coordinates from 14,371 published studies and 3,228 semantic terms extracted from study abstracts. Abstract retrieval and preprocessing were conducted using built-in NiMARE functions with article abstracts obtained from PubMed.

A GC-LDA model was trained on a subset of semantic terms derived from the term classifications defined in Peraza et al. (2024), excluding terms labeled as “Non-specific,” yielding a subset of 1395 terms. The GC-LDA framework jointly models spatial activation patterns and semantic term distributions to estimate topic maps that maximize correspondence between functional terminology and brain activity. From this model, voxelwise posterior probability maps *p(topic|voxel)*, representing the probability of each topic at a given voxel, were retained. Using the selected semantic term subset, 200 GC-LDA topic meta-analytic maps were generated.

For functional decoding, we computed spatial correlations between unthresholded ALE Z-valued maps and the full set of GC-LDA topic maps. For the disease-specific hyper- and hypometabolism analyses, which have unidirectional effects, the 10 topics demonstrating the strongest positive correlations with each ALE map were retained. For the subtraction analyses, which produce signed maps reflecting differences between diseases, the 10 topics with the strongest positive correlations and the 10 topics with the strongest negative correlations for each contrast were retained. Positive correlations indicate greater correspondence with regions more strongly represented in the first disease of the contrast, whereas negative correlations indicate correspondence with regions more strongly represented in the second disease. For each retained topic, the three highest-weighted associated terms were identified, and only terms classified as “Functional” were reported.

GC-LDA decoding identifies spatial correspondence between ALE-derived metabolic patterns and functional terminology derived from task-based neuroimaging studies of largely healthy populations in the Neurosynth database. These correspondences reflect the degree to which the spatial distribution of metabolic patterns overlaps with literature-derived activation patterns. It does not demonstrate that the hypo- and hypermetabolic patterns cause, co-occur with, or are functionally equivalent to associated cognitive or behavioral processes in the disease populations studied here. Decoding results are therefore hypothesis-generating in nature and require validation in studies correlating regional metabolism with behavioral performance in these populations.

#### Sensitivity analyses

We evaluated cluster robustness in the primary analyses using jackknife, focus counter, fail-safe N[50], and resampled stability analyses[51]. The jackknife analysis quantified the proportional change in each cluster statistic after removal of each experiment for which we report the mean contribution across all experiments and the maximum contribution from the most influential experiment. The focus counter summarized the number of experiments and reported foci contributing to each cluster and we report the number and proportion of experiments contributing at least one focus, and the total number of foci within each cluster. Fail-safe N analyses added simulated null experiments generated from the observed distributions of sample sizes and focus counts, with null foci randomly sampled within the analysis mask. We implemented this with 10 independent null-study sets (drawers) evaluated using binary search capped at 5 × k null studies and 5,000 Monte Carlo iterations per evaluated level. We report the median fail-safe N with 2.5th–97.5th percentile range across drawers, and the number of right-censored drawers in which the cluster remained significant at the search limit. Resampled stability quantified voxelwise reproducibility across subsets containing approximately 75% of the experiments (1,000 random subsets, or exhaustive resampling when fewer than 1000 unique subsets existed). We report the mean voxelwise stability and the proportion of cluster voxels reaching stability thresholds of 0.50 and 0.80. These results are presented in Supplemental Table 3 and include spatial characteristics (volume, peak Z-statistic, MNI coordinates, and anatomical labels) alongside robustness summaries for each diagnostic.

#### Spatial representations

All results were plotted with *nilearn* (v 0.13.0)[52] and significant clusters were anatomically localized using the Automated Anatomical Labeling (AAL), Harvard-Oxford, and Juelich atlases implemented in *atlasreader* (v 0.3.2)[53]. Identified clusters from the various analyses were set to their maximum familywise error corrected Z-value by NiMARE. Thus, cluster coordinates define the cluster center of mass rather than peak intensity coordinates. Given specific atlas definitions, it is possible for the center of mass to occur outside of a defined atlas region or to be localized to gray matter in one atlas and white matter in another.

## Results

Search of literature resulted in retrieval of 21,080 records, of which 10,863 duplicates and an additional 2943 were removed for other reasons (conference abstracts, comments, editorials, etc.) (see Figure 1, Supplemental Table 1). Title and abstract screening removed another 4728 records. Full text articles were reviewed for 2,074 records using the inclusion criteria. A total of 130 records were included in the analysis[29,32–36,54–177]. Table 1 provides aggregated details on disease-specific breakdowns and Supplemental Table 2 provides study-level details.

**Figure 1.**
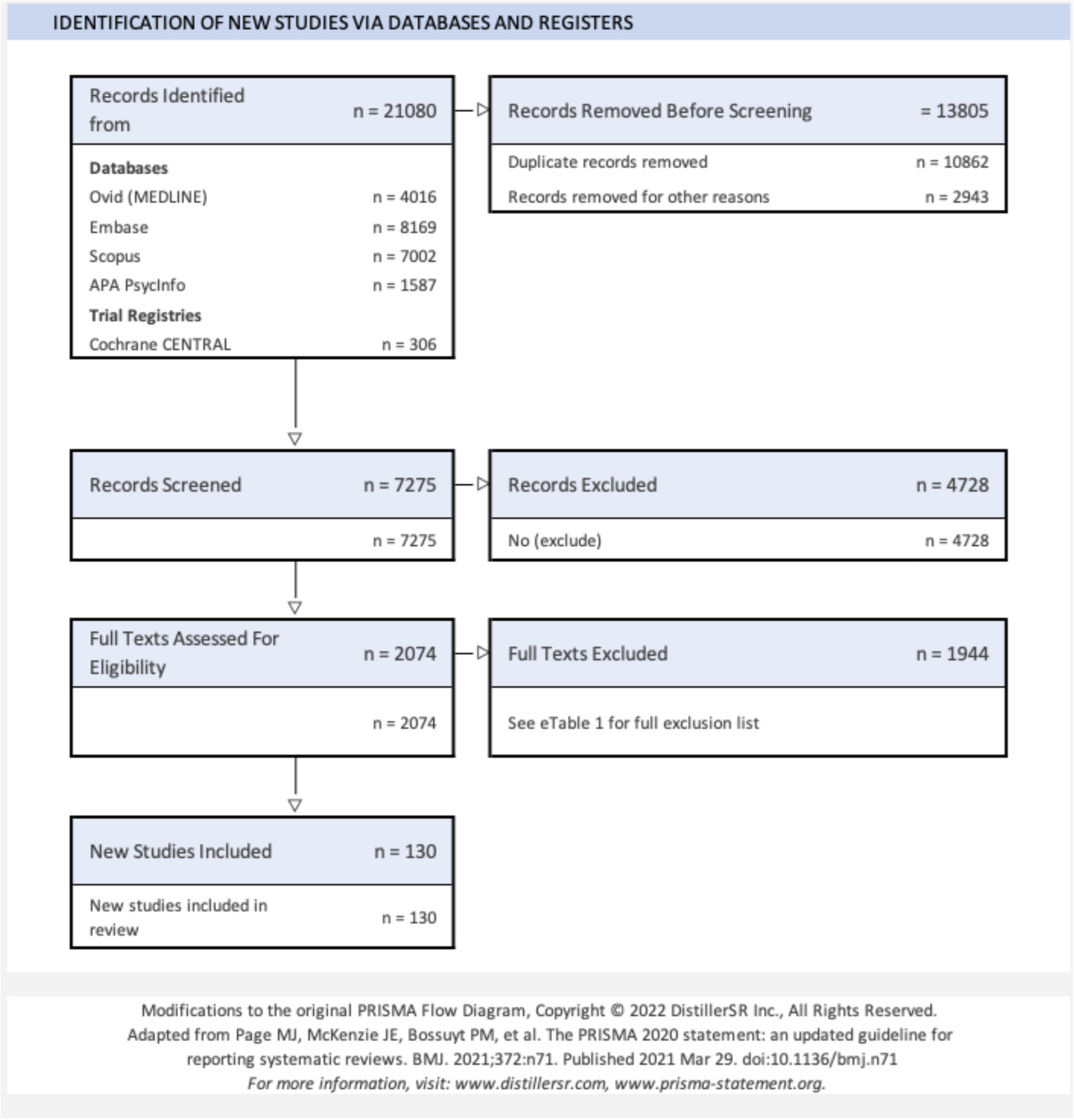
PRISMA flow chart of literature search and selection criteria for studies included in the meta-analysis.

**Table 1.** Summary of included studies by group.

| Disease group | Metabolic comparison | Number of studies reporting | Number of groupwise comparisons | No. of reported coordinates | Control sample size | Control age (mean± SD) | A<sup>2</sup>ND sample size | A<sup>2</sup>ND age (mean± SD) |
| --- | --- | --- | --- | --- | --- | --- | --- | --- |
| A<sup>2</sup>ND | Disease < Control | 129[29,32–36,54–134,136–177] | 171 | 1382 | 3499 | 65.7±5.96 | 5298 | 68.0±5.60 |
|  | Disease > Control | 34[29,32,33,35,36,56,59,66,69,81,83,85,86,103,108,112,116,120,123,125,126,131–133,135–137,144,146,155,157,163,165,176] | 42 | 261 | 956 | 62.1±5.64 | 1504 | 64.2±5.50 |
| AD | Disease < Control | 84[29,55,58,60,67–69,71,74–79,86,89,90,95–99,102,106,107,110,111,116,118,119,121,122,128,129,133,139–143,145,148–150,152,153,164,165,170] | 104 | 807 | 2213 | 67.2±6.11 | 2885 | 69.9±5.28 |
|  | Disease > Control | 10[33,35,36,69,103,108,116,120,133,165] | 10 | 66 | 196 | 66.2±7.03 | 235 | 69.7±5.53 |
| PD | Disease < Control | 31[33,34,36,56,57,81,82,86,91,93,100,112–114,117,123,125,126,129,132,144,147,151,155–157,166,167,172,175,176] | 45 | 326 | 592 | 62.8±5.07 | 966 | 65.8±4.96 |
|  | Disease > Control | 15[33,36,56,81,86,112,123,125,126,132,135,144,155,157,176] | 20 | 142 | 319 | 59.8±5.32 | 491 | 62.6±4.75 |
| ALS | Disease < Control | 17[32,59,61–66,83,85,88,131,136,137,146,161,163] | 22 | 249 | 807 | 62.5±2.84 | 1387 | 62.9±3.17 |
|  | Disease > Control | 10[32,59,66,83,85,131,136,137,146,163] | 12 | 53 | 459 | 61.3±2.40 | 718 | 62.0±3.37 |

### Disease-specific analyses

Disease-specific ALEs revealed distinct metabolic patterns (Figure 2A-C, Table 2, Supplemental Table 3). AD was primarily characterized by extensive hypometabolism localized to 8 significant clusters across the precuneus and bilateral angular gyri, alongside 3 significant hypermetabolic clusters in the postcentral gyrus as well as the right putamen and left thalamus. ALE analyses revealed 7 significant hypometabolic and 5 significant hypermetabolic clusters in PD. Hypometabolic clusters were primarily located in the bilateral angular gyrus, and striatum, while hypermetabolic clusters were identified in the cerebellum, left paracentral lobule, bilateral thalami, and right gyrus rectus. In ALS, 7 clusters were identified as hypometabolic, primarily in the middle and inferior frontal regions along with the precentral gyrus. Hypermetabolism in ALS brain was detected in two clusters, one in the hippocampus and one in the brainstem.

**Figure 2.**
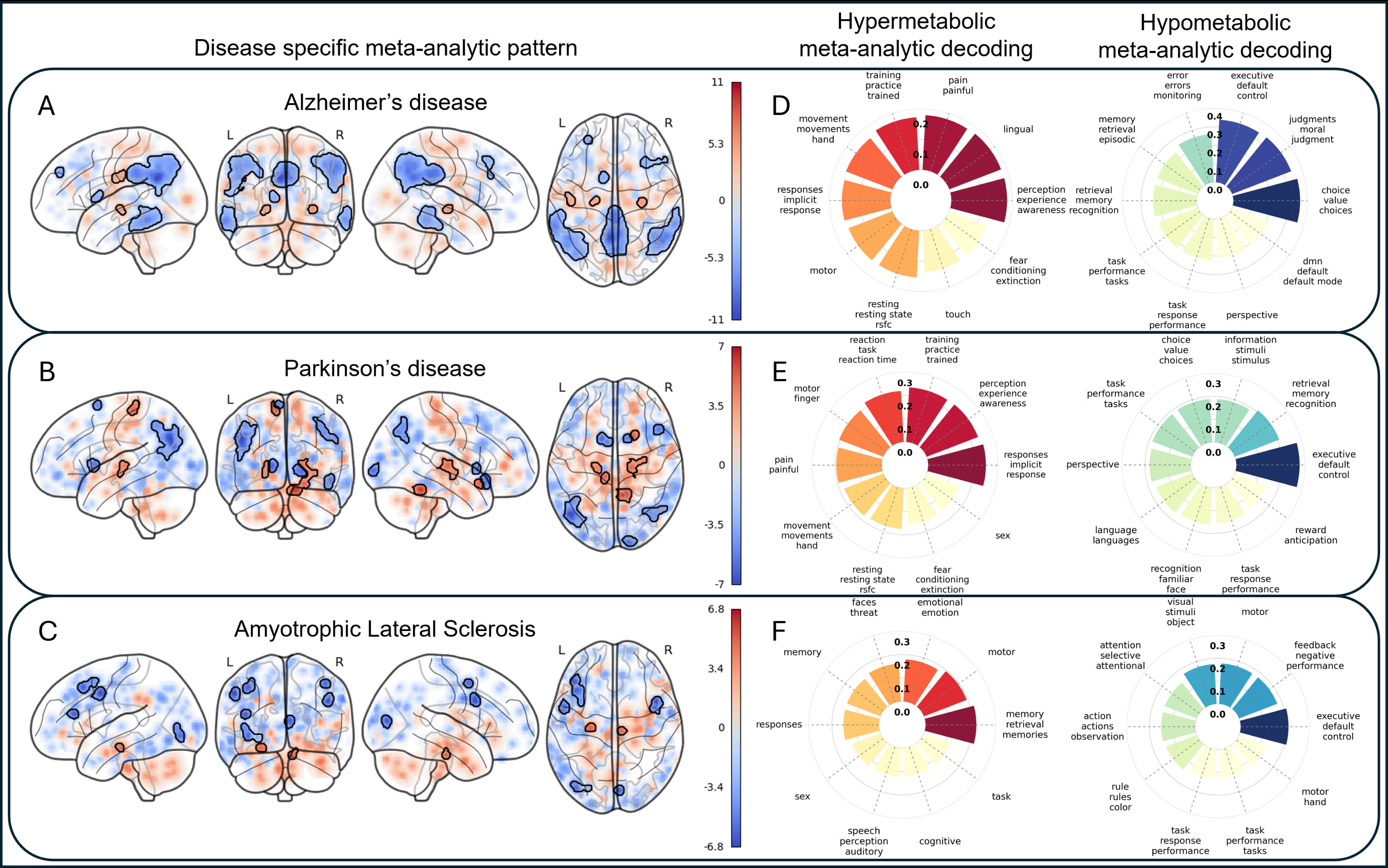
Disease-specifc metabolic spatial patterns spatially align with clinically relevant functional decoding terms. Z-score maps for disease-specific ALE outcomes are illustrated on a glass brain for Alzheimer’s disease (A), Parkinson’s disease (B) and amyotrophic lateral sclerosis (C). Uncorrected outcomes are presented transparently while statistically significant (p < 0.001, FWE-corrected p < 0.05) hyper- (positive values, red colors) and hypometabolism (negative values, blue colors) are presented opaquely and outlined in black. Radar plots indicate correlation values from functional meta-analytic decoding of the unthresholded Z-score maps for Alzheimer’s disease (D), Parkinson’s disease (E), and amyotrophic lateral sclerosis (F). Positive associations (greater association with hypermetabolic values) are presented in red and negative associations (greater association with hypometabolic values) are presented in blue. All correlation values are reported with absolute *r* values for ordering on radar plots.

**Table 2.**
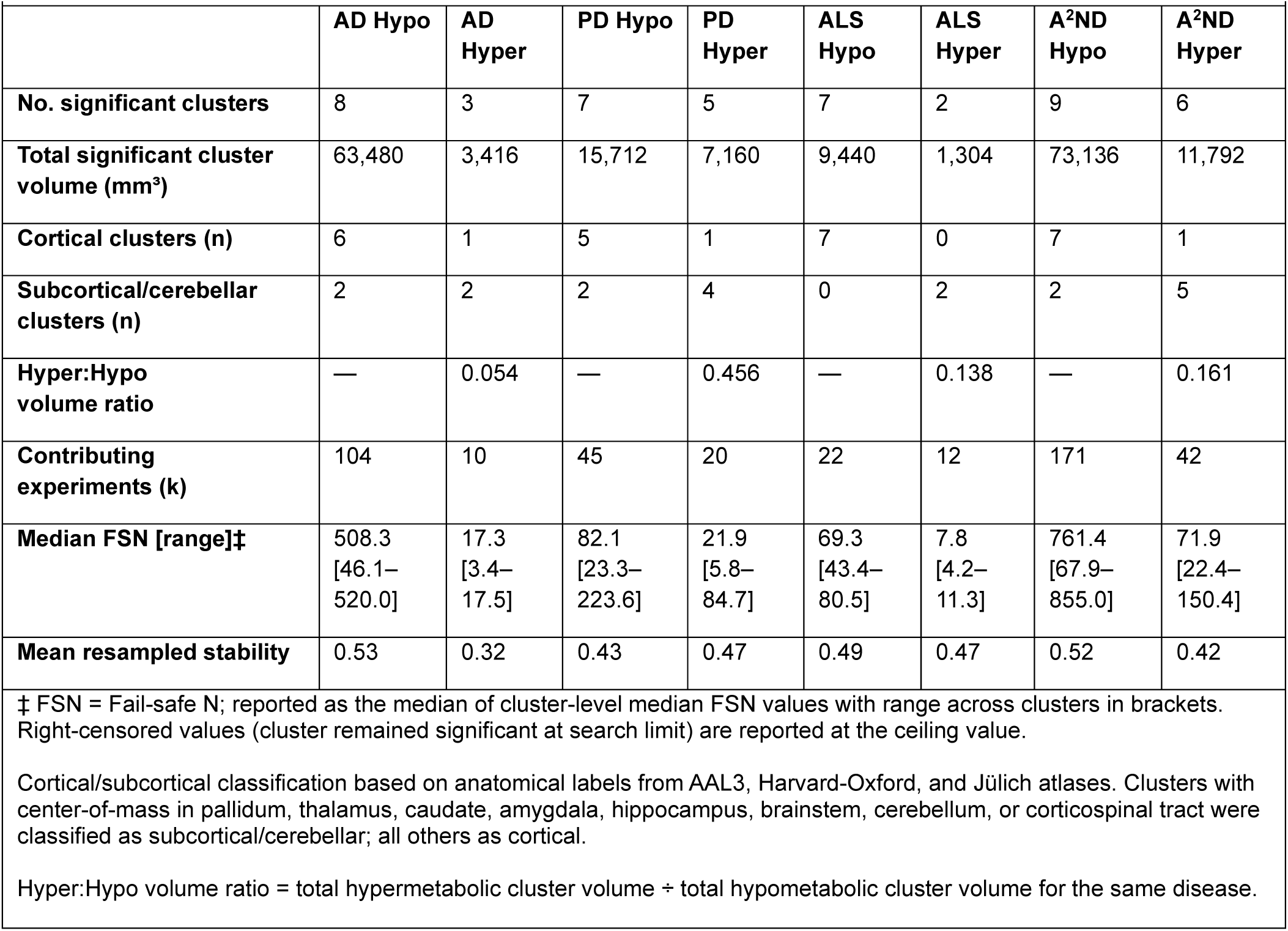
Quantitative summary of disease-specific and A^2^ND meta-analytic results.

Meta-analytic functional decoding across the three diseases (Figure 2D-F) identified spatial correspondence between hypometabolic patterns and functional terminology from the neuroimaging literature. Hypometabolic patterns had the strongest spatial correspondence with terms related to cognitive functions, particularly executive control, task performance and monitoring, decision-making processes, and default mode function. These spatial correspondences exhibited distinct profiles (e.g., choices and value judgments in AD, stimulus and task monitoring in PD, and visual attention in ALS), while converging into higher-order cognitive function associations. By contrast, hypermetabolism functional associations were more disease specific. Hypermetabolism in AD and PD corresponded most strongly to terms related to general perception, pain, and movement, whereas ALS hypermetabolism corresponded most strongly to memory-related terms.

Table 2 provides a quantitative summary of meta-analytic outcomes across diseases. Several patterns are evident: (1) hypometabolism is consistently more spatially extensive than hypermetabolism across all diseases, with hyper:hypo volume ratios ranging from 0.054 in AD to 0.456 in PD; (2) hypometabolic clusters are predominantly cortical while hypermetabolic clusters are predominantly subcortical or cerebellar; (3) ALS exhibits exclusively cortical hypometabolism and exclusively subcortical hypermetabolism, representing the most extreme anatomical dissociation among the three diseases; and (4) robustness metrics (fail-safe N and resampled stability) scale with the size of the contributing literature, with hypometabolic patterns in AD and the combined A^2^ND analysis exhibiting the strongest support for robust clusters (see Sensitivity Analyses below and Supplemental Table 3 for full details).

### Conjunction analyses

Common hypometabolic outcomes, revealed through conjunction analyses, were limited in both frequency and spatial extent (Supplemental Table 4). There were no common statistically significant hypo- or hypermetabolic clusters across all three diseases. Only Alzheimer’s and Parkinson’s exhibited any shared significance with four hypometabolic clusters (bilateral angular gyri, left caudate, left middle temporal gyrus) and two hypermetabolic clusters (left thalamus and right putamen; Supplemental Table 4).

### Subtraction analyses

Pairwise subtraction analyses highlighted disease specific metabolic outcomes (Figure 3A-C). Greater evidence of hypometabolism in AD compared to either PD or ALS corresponded spatially to regions typically engaged during cognitive functions, including memory, attention, and emotion-related processing. In PD, regions identified as hypometabolic compared to AD corresponded to functional patterns associated with information, stimulus, and response processing while hypometabolism compared to ALS showed greatest correspondence with functional terms related to risk/reward systems as well as interpersonal cognition. ALS hypometabolism compared to either AD or PD corresponded more strongly with functional profiles of motor function. The spatial extent and location of identified clusters is presented in Supplemental Table 5.

**Figure 3.**
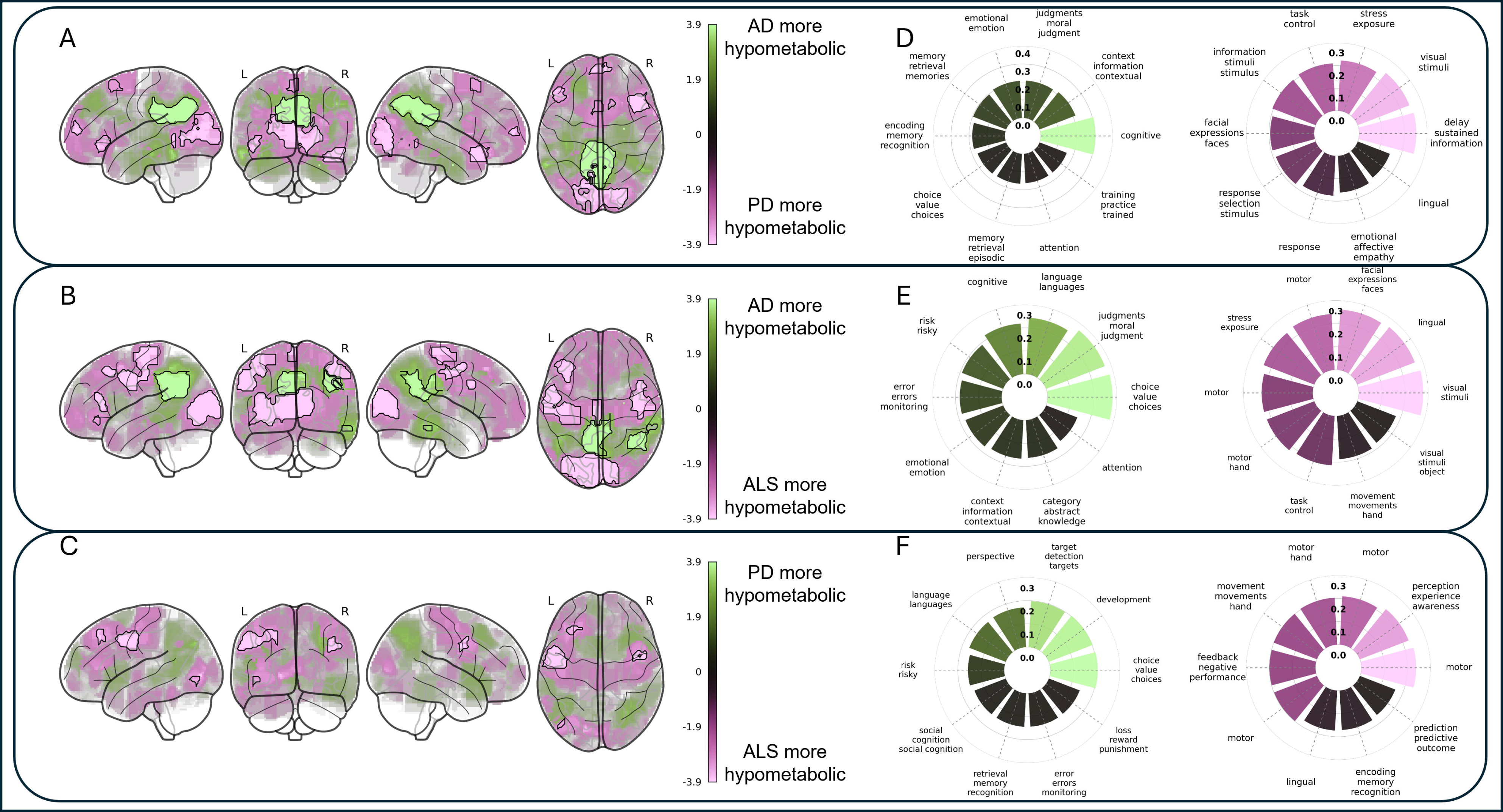
Differences in pairwise disease-specific metabolic spatial patterns correspond to functional phenotypes. Z-score maps for ALE subtraction analyses are illustrated on a glass brain for Alzheimer’s – Parkinson’s disease (A), Alzheimer’s disease – amyotrophic lateral sclerosis (B) and Parkinson’s disease – amyotrophic lateral sclerosis (C). Uncorrected outcomes are presented transparently while statistically significant (*p* < 0.001, and a cluster extent of at least 25 connected voxels (200mm^3^)) are presented opaquely and outlined in black. Positive (green) and negative (purple) values encode which disease group had larger values. Radar plots indicate correlation values from functional meta-analytic decoding of the unthresholded ALE subtraction Z-score maps for each disease pair contrast (D-F). Positive associations are presented in green and negative associations are presented in purples, aligning with contrast directionality in A-C. All correlation values are reported with absolute *r* values for ordering on radar plots.

Greater hypermetabolism was detected in PD compared to ALS in one cluster localized to the left paracentral lobule and primary motor cortex. No other hypermetabolic differences met the statistical threshold for significance.

### A^2^ND transdiagnostic metabolic disruption

Combining all three diseases together revealed large and spatially diffuse patterns of hypometabolism and smaller, focal areas of hypermetabolism (Figure 4A). Hypometabolism was identified in 9 significant clusters, the largest of which overlapped with the bilateral angular gyri, precuneus, and both right and middle temporal gyri (Figure 4B, Supplemental Table 3). Hypermetabolism was identified in 6 statistically significant clusters, the largest of which were located over the bilateral thalami, right hippocampus and amygdala and cerebellum (Figure 4C, Supplemental Table 3). Functional decoding revealed that hypermetabolic patterns showed strongest correspondence with terms related to sensory perception, motor performance, and emotional responses (e.g., fear) whereas hypometabolic patterns exhibited strongest correspondence with terms related to cognitive performance in choices, judgments, error monitoring, memory, language, and multiple networks including the executive, default, and fronto-parietal control networks overall (Figure 4D-E).

**Figure 4.**
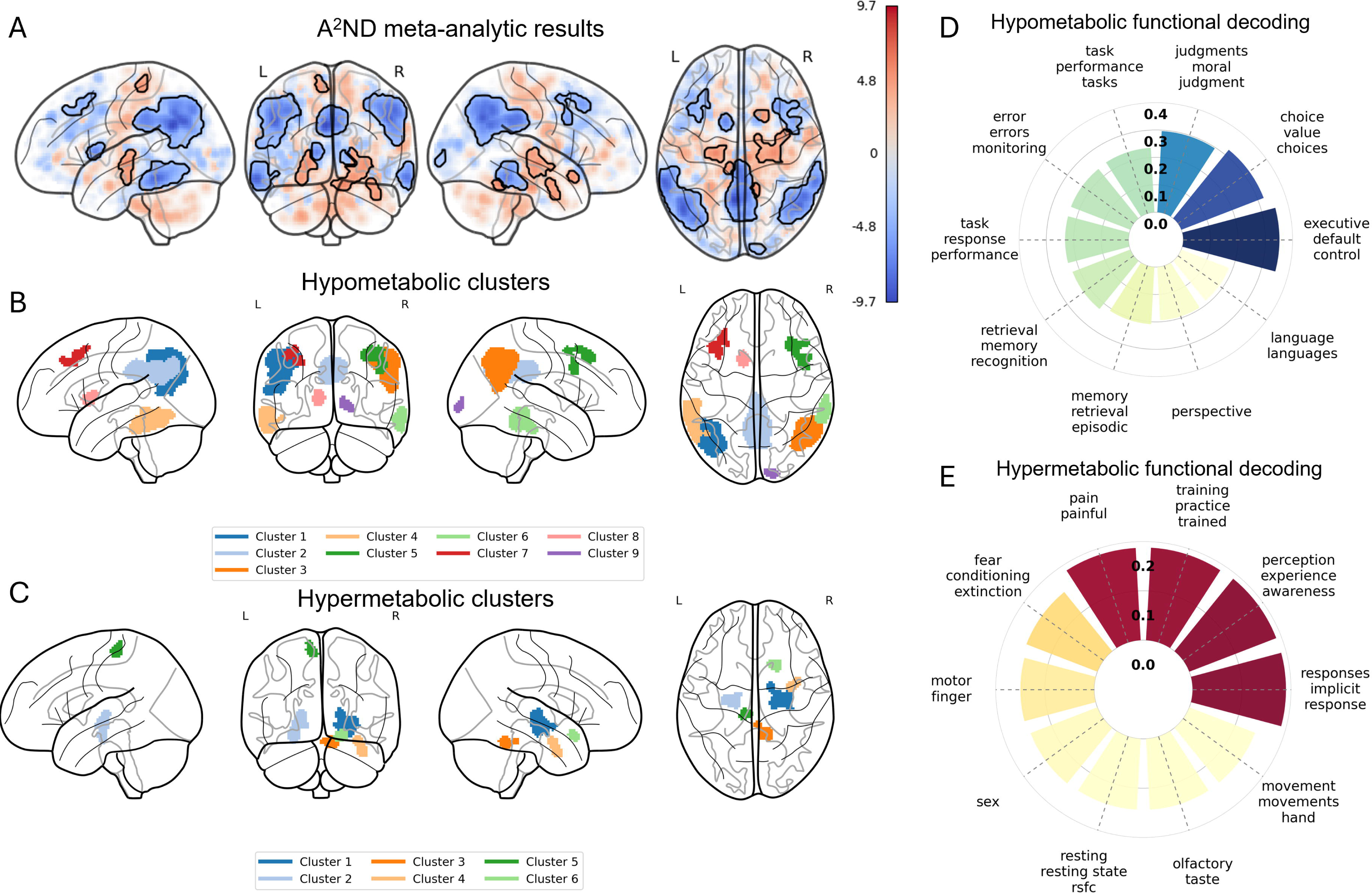
Transdiagnostic metabolic outcomes link hypo- and hypermetabolism with cognitive, sensory, and emotional functional decoding. Transdiagnostic ALE meta-analysis Z-scores (A; red = hypermetabolism, blue = hypometabolism) as well as hypo- (B) and hypermetabolic (C) cluster maps overlaid on glass brains reveal widespread hypometabolism and focal hypermetabolism as a common features of age-associated neurodegenerative disease. Radar plots indicate correlation values from functional meta-analytic decoding of the unthresholded Z-score maps. Negative associations (D) connect hypometabolic patterns with executive and control network cognitive functions. Positive associations (E) connect hypermetabolism with sensory and emotional function. All correlation values are reported with absolute *r* values for ordering on radar plots.

## Sensitivity analyses

Sensitivity analyses revealed a gradient of evidential strength that aligned with the size of the contributing literature and the metabolic direction of the contrast (see Supplemental Table 3). The hypometabolism analyses, particularly the A^2^ND (k = 171) and AD (k = 104), had the most robust clusters. The largest clusters reached the fail-safe N search ceiling (medians of 855 and 520, right-censored across all 10 drawers), achieved mean resampled stability of 0.65–0.77 with 41–64% of voxels exceeding the 0.80 stability threshold, were supported by 30–72% of included experiments, and showed minimal jackknife dependence (maximum contributions ≤ 0.05). Smaller hypometabolic clusters in these analyses (including frontal and occipital regions) exhibited weaker profiles, with stability below 0.40, no voxels at the 0.80 threshold, and support from fewer than 10% of experiments, though fail-safe N values (46–134) still indicated moderate resilience. PD hypometabolism (k = 45) fell between these extremes, with the largest cluster (left angular/inferior parietal) reaching a fail-safe N of 224 (right-censored in 9/10 drawers) and stability of 0.53, whereas ALS hypometabolism (k = 22) produced clusters with moderate stability (0.37–0.60) and fail-safe N values of 43–81.

Hypermetabolism analyses consistently revealed weaker robustness, reflecting smaller contributing literature bases. A^2^ND (k = 42) and PD (k = 20) hypermetabolism clusters achieved fail-safe N values of 22–150 and stability of 0.35–0.62. AD hypermetabolism (k = 10) and ALS hypermetabolism (k = 12) yielded the least robust clusters, with stability ranging from 0.29–0.55, fail-safe N values as low as 3.4– 4.2, support from as few as 2–3 experiments, and jackknife maximum contributions up to 0.57, indicating substantial dependence on individual studies. Across all analyses, larger clusters with higher peak statistics generally showed stronger robustness profiles, and jackknife mean contributions decreased systematically from approximately 0.10 (k = 10) to 0.006 (k = 171), consistent with expected dilution of individual experiment influence in larger study sets.

## Discussion

Outcomes of the meta-analysis reported herein provide insights into the spatial patterns of altered glucose metabolism across AD, PD, and ALS. The transdiagnostic meta-analysis, supported by the disease-specific meta-analyses, revealed both hyper- and hypometabolic responses occur across AD, PD and ALS with distinct disease-specific patterns. The conjunction analyses demonstrate that there is not a singular unifying transdiagnostic metabolic spatial pattern defined by jointly expressed metabolic dysregulation. Rather, the data indicate that i) both hyper- and hypometabolism are observed in each disease; ii) there are statistically robust disease-distinguishing patterns of hypometabolism; iii) each disease expresses a unique pattern of hypermetabolic regions; and iv) meta-analytic decoding of these differences support distinct clinically observed behavioral and functional disease related outcomes.

The transdiagnostic meta-analysis, pooled across the diseases, identified hypometabolism predominantly in posterior cortical regions, specifically the posterior cingulate, angular gyri, and precuneus, with additional involvement of fronto-parietal regions. These regions are hubs within the default mode and control networks. Meta-analytic decoding indicated that the spatial distribution of transdiagnostic hypometabolism corresponds to regions typically involved in executive control, cognitive performance monitoring, and decision-making tasks in the healthy neuroimaging literature. Prior findings demonstrate high metabolic demands from the default mode network at rest[178] and a high reliance on aerobic glycolysis to support network suppression during internal (i.e., working memory) tasks compared to external tasks (i.e. vigilant attention)[179]. Reduced glucose uptake due to neurodegenerative disease, as identified here, overlaps spatially with regions that support cognitive performance in healthy populations, consistent with a link between metabolic decline and clinically observed cognitive deficits in memory, task performance, and judgment across age-associated neurodegenerative disease.

Hypermetabolism was less spatially extensive than hypometabolism and primarily localized to subcortical and limbic regions, including the hippocampus and thalamus, as well as the cerebellum. Meta-analytic decoding indicated that the spatial distribution of transdiagnostic hypermetabolism corresponded most strongly to functional terms related to sensory and perceptual outcomes, consistent with past reports of heightened pain perception[180,181] as well as fear and anxiety in neurodegenerative disease[182–184]. Unexpectedly, the hypermetabolic pattern showed spatial correspondence with terms related to hand and finger motor outcomes, despite general motor impairments in both PD[185] and ALS[186]. The anatomical localization of these findings together with the disease-specific localization of hypermetabolic regions in structures with known functional connectivity to affected cortical networks (e.g., cerebellum in PD, hippocampus in ALS), suggests that increased regional glucose uptake may plausibly reflect a compensatory response to cortical metabolic hypometabolism. However, FDG-PET does not provide cellular specificity, and alternative mechanisms such as general metabolic insufficiency, neuroinflammation, and gliosis cannot be excluded without multimodal imaging approaches offering cellular-level resolution[187,188]. Distinguishing among these possibilities will require studies combining FDG-PET with markers of neuroinflammation (e.g., TSPO-PET), synaptic density, and direct behavioral-metabolic correlations in these populations.

Conjunction analyses revealed limited spatial overlap in hyper- or hypometabolism across diseases. Rather, the spatial distribution of metabolic deficits is consistent with disease-specific neurodegenerative functional regions. AD-related hypometabolism was centered in the precuneus and bilateral angular gyri, regions implicated in amyloid deposition and tau pathology[189–191], whereas hypermetabolism occurred in the postcentral gyrus as well as thalamus and putamen. In PD, occipital and caudate hypometabolism, along with cerebellar hypermetabolism, aligns with reports of motor[192] and visual processing dysfunction[193] and compensatory basal ganglia-cerebellar interactions[194,195]. In ALS, inferior frontal and occipital hypometabolism, alongside hippocampal hypermetabolism, likely reflects the combined effects of motor network degeneration and frontotemporal involvement[196–198]. Meta-analytic decoding of hypometabolism subtraction analyses revealed spatial correspondences that parallel these clinical distinctions, though these associations remain hypothesis-generating. Compared to either PD or ALS, AD-related hypometabolism corresponded to terms related to cognition, memory, decision making, and emotion-related processes. PD-specific hypometabolism showed greatest correspondence with functional terms reflecting stimulus processing, responsiveness, and decision-making behaviors related to both risk and reward. ALS-specific hypometabolism primarily showed greater correspondence to functional terms reflecting motor function compared to either AD or PD, reflecting the more unique nature of this motor neuron disease, with additional associations to memory and perception function compared to PD. Collectively, these findings suggest that while bioenergetic dysfunction is a shared feature of neurodegeneration, each disease follows a distinct metabolic distribution that reflects its clinical and functional profile.

### Implications for treatment

The disease-specific patterns of hypo- and hypermetabolism identified in this meta-analysis indicate that metabolic dysregulation could impact A^2^ND functional recovery and therapeutic responsiveness. Hypometabolism identified here parallels the general clinical presentations of these diseases, suggesting that impaired bioenergetic function likely limits the extent to which these systems can return to more normal function, even when core upstream pathological processes are addressed. Recent work within AD highlights the fact that individuals can remain cognitively unimpaired even with high amyloid and tau burden, while those with metabolic deficits are more likely to progress to clinical AD diagnoses[199–202]. It follows that metabolic dysregulation is likely a key driver of clinical presentation and therefore a critical target for therapeutic engagement, rather than solely a downstream disease biomarker. If bioenergetic preservation is key to sustaining function despite pathology, then this shifts the paradigm from exclusively targeting risk factors or pathology drivers to keeping people on the “right side of normal” earlier rather than later. Surveillance shifts from symptom presentation after it’s too late to early multimorbidity risk-informed monitoring that drives intervention when in-brain bioenergetic shifts are small[203]. This facilitates leveraging therapeutics that address bioenergetic deficits coupled with pathology-modifying or regenerative approaches to sustain, preserve, and recover function[204–206].

### Limitations

A critical observation is the paucity of studies identifying hypermetabolism across any of the diseases. Hypermetabolic outcomes across the A^2^ND groups were reported in only 34 out of the 130 studies included here (Table 1). One essential consideration is the choice of reference region on subsequent case-control comparisons. Past work indicates that choice of reference region significantly impacts the identification and interpretation of both hypo- and hypermetabolism[207]. We did not control for nor restrict reference region definitions and acknowledge that this may have an unquantifiable impact on the outcomes reported in the source literature and identified in the present analyses. A further possible explanation for this is an intentional emphasis on hypometabolism while ignoring hypermetabolism.

While some studies explicitly identified a lack of hypermetabolic responses meeting study-specific criteria for statistical significance or reporting[78,79,124], others specifically describe only considering the hypometabolic response[34,177] and others further neglect reporting clearly either in methods or results whether the hypermetabolic response was considered[77,93].

Further, the included studies span nearly 40 years (1998-2025), during which time technological innovations have improved FDG-PET data acquisition and quality, as well as analytic capabilities and best practices. Disease diagnostic guidelines have evolved in these 40 years, particularly for Alzheimer’s disease, as well. The consequence of this is that these analyses incorporate a diverse range of disease definitions, spatial resolution in the source data, and processing choices at the study level (e.g., normalization, SUV reference region, covariate inclusion, and cluster forming definitions) all of which covary across studies and publication eras. A study using pons normalization, for example, also reflects a particular set of software, covariate, threshold, and diagnostic era choices. Stratified subgroup analyses by any single methodological factor confounds it with all co-occurring features, precluding interpretable subgroup contrasts. The sensitivity analyses presented here (jackknife, fail-safe N, focus counter, resampled stability) address a complementary question: whether findings are driven by individual studies or are resilient to study-set perturbation. These diagnostics demonstrate that hypometabolic findings are robust to individual study influence, but they cannot isolate contributions of specific methodological choices. This limitation applies particularly to hypermetabolism findings, which are supported by fewer studies and show greater individual-study dependence.

Finally, the ALE subtraction analyses identifying disease-specific differences in metabolic patterns relied on an uncorrected voxel-level threshold (*p* < 0.001) combined with a minimum cluster extent (200 mm³), as no well-validated multiple comparisons correction currently exists for this procedure within the ALE framework. While the permutation-based null distribution (10,000 iterations) provides a non-parametric basis for significance testing at the voxel level, the absence of cluster-level FWE correction increases the risk of false positive findings compared to the primary ALE and conjunction analyses.

Disease-specific subtraction results should therefore be considered more exploratory than the primary and conjunction analyses. However, the spatial patterns identified are consistent with known clinical phenotypes – such as AD-specific hypometabolism in posterior cingulate/precuneus regions and ALS-specific hypometabolism in motor cortex – which provides face validity. Independent replication with formal correction methods, as they become available, would strengthen confidence in these disease-distinguishing patterns

## Conclusions

Glucose dysregulation in age-associated neurodegenerative disease includes both hypo- and hypermetabolic responses, in disease specific spatial distributions. Hypometabolic patterns are spatially consistent with the clinical presentations of these diseases, and meta-analytic decoding reveals spatial correspondence between regions of metabolic decline and functional terms associated with the cognitive, motor, and behavioral domains most affected in each disease. The consistent identification of hypermetabolic regions across all three diseases, predominantly in subcortical and cerebellar structures anatomically distinct from hypometabolic cortical regions, raises the possibility that increased regional glucose uptake reflects a compensatory response to cortical metabolic failure. This interpretation is supported by the spatial complementarity of hyper- and hypometabolic patterns and by the disease-specific localization of hypermetabolism in regions with known functional relationships to affected cortical networks (e.g., cerebellum in PD, hippocampus in ALS). However, FDG-PET alone cannot distinguish this from other possible mechanisms, such as neuroinflammation or methodological considerations, and future research should prioritize determining the biological basis of regional hypermetabolism. If compensatory adaptation is confirmed, this would have direct implications for therapeutic strategies, suggesting that metabolic stabilization should be initiated early, before compensatory capacity is exhausted, and coupled with disease-modifying or regenerative approaches to sustain and recover function.

## Data Availability

All data generated or analysed during this study are included in this published article and its supplementary information files.

## Acknowledgements

We acknowledge Amanda Tack for her contributions in article screening and project management.

