## Supplemental for "Meta-analysis of glucose metabolism across Alzheimer’s, Parkinson’s and ALS reveals distinct patterns of cortical hypometabolism versus subcortical hypermetabolism with disease-specific spatial signatures"

**Supplement A**

**Full search strategies**

**Ovid MEDLINE(R) ALL**

| | |
|---|---|
| 1 | Fluorodeoxyglucose F18/ |
| 2 | glucose/ |
| 3 | Fluorodeoxyglucose.ti,ab,kf. |
| 4 | glucose.ti,ab,kf. |
| 5 | 18fluorodeoxyglucose.ti,ab,kf. |
| 6 | FluorodeoxyglucoseF18.ti,ab,kf. |
| 7 | (fluoro adj2 deoxy adj2 glucose).ti,ab,kf. |
| 8 | FDG.ti,ab,kf. |
| 9 | Deoxyfluoroglucose.ti,ab,kf. |
| 10 | fludeoxyglucose.ti,ab,kf. |
| 11 | deoxyglucose.ti,ab,kf. |
| 12 | glucotrace.ti,ab,kf. |
| 13 | steripet.ti,ab,kf. |
| 14 | (0Z5B2CJX4D or Fluorodeoxyglucose or 63503-12-8).rn. |
| 15 | or/1-14 |
| 16 | Positron-Emission Tomography/ |
| 17 | "Positron Emission Tomography".ti,ab,kf. |
| 18 | (Positron adj2 Emission adj2 Tomography).ti,ab,kf. |
| 19 | PET.ti,ab,kf. |
| 20 | or/16-19 |
| 21 | Amyotrophic Lateral Sclerosis/ |
| 22 | (Amyotrophic adj2 Lateral adj2 Sclerosis).ti,ab,kf. |
| 23 | "lou gehrig* disease".ti,ab,kf. |
| 24 | "guam disease".ti,ab,kf. |
| 25 | or/21-24 |
| 26 | 15 and 20 and 25 |
| 27 | Parkinson Disease/ |
| 28 | "Parkinson* Disease".ti,ab,kf. |
| 29 | "paralysis agitans".ti,ab,kf. |
| 30 | "primary parkinsonism".ti,ab,kf. |
| 31 | or/27-30 |
| 32 | 15 and 20 and 31 |
| 33 | multiple sclerosis/ or multiple sclerosis, chronic progressive/ or multiple sclerosis, relapsing-remitting/ |
| 34 | ((multiple or disseminated) adj2 sclerosis).ti,ab,kf. |
| 35 | or/33-34 |
| 36 | 15 and 20 and 35 |
| 37 | Alzheimer Disease/ |

| | |
|---|---|
| 38 | "Alzheimer* Disease".ti,ab,kf. |
| 39 | (dementia adj2 (presenile or senile or alzheimer* or degenerative)).ti,ab,kf. |
| 40 | (alzheimer* adj (sclerosis or syndrome)).ti,ab,kf. |
| 41 | "primary senile".ti,ab,kf. |
| 42 | or/37-41 |
| 43 | 15 and 20 and 42 |
| 44 | 26 or 32 or 36 or 43 |
| 45 | Limit 44 to English language |

**Embase.com**

| No. | Query |
|---|---|
| #37 | (#18 OR #24 OR #28 OR #35) AND [english]/lim |
| #36 | #18 OR #24 OR #28 OR #35 |
| #35 | #12 AND #34 |
| #34 | #29 OR #30 OR #31 OR #32 OR #33 |
| #33 | 'primary senile':ti,ab,kw |
| #32 | (alzheimer* NEAR/2 (sclerosis OR syndrome)):ti,ab,kw |
| #31 | (dementia NEAR/2 (presenile OR senile OR alzheimer* OR degenerative)):ti,ab,kw |
| #30 | 'alzheimer* disease':ti,ab,kw |
| #29 | 'alzheimer disease'/de |
| #28 | #12 AND #27 |
| #27 | #25 OR #26 |
| #26 | ((multiple OR disseminated) NEAR/2 sclerosis):ti,ab,kw |
| #25 | 'multiple sclerosis'/de OR 'progressive multiple sclerosis'/de OR 'relapsing remitting multiple sclerosis'/de OR 'tumefactive multiple sclerosis'/de |
| #24 | #12 AND #23 |
| #23 | #19 OR #20 OR #21 OR #22 |

| | |
|---|---|
| #22 | 'primary parkinsonism':ti,ab,kw |
| #21 | 'paralysis agitans':ti,ab,kw |
| #20 | 'parkinson* disease':ti,ab,kw |
| #19 | 'parkinson disease'/de |
| #18 | #12 AND #17 |
| #17 | #13 OR #14 OR #15 OR #16 |
| #16 | 'guam disease':ti,ab,kw |
| #15 | 'lou gehrig* disease':ti,ab,kw |
| #14 | (amyotrophic NEAR/2 lateral NEAR/2 sclerosis):ti,ab,kw |
| #13 | 'amyotrophic lateral sclerosis'/de |
| #12 | #7 AND #11 |
| #11 | #8 OR #9 OR #10 |
| #10 | pet:ti,ab,kw |
| #9 | (positron NEAR/2 emission NEAR/2 tomography):ti,ab,kw |
| #8 | 'positron emission tomography'/de |
| #7 | #1 OR #2 OR #3 OR #4 OR #5 OR #6 |
| #6 | 'glucose'/de |
| #5 | glucotrace:ti,ab,kw OR steripet:ti,ab,kw |
| #4 | (fluoro NEAR/2 deoxy NEAR/2 glucose):ti,ab,kw |
| #3 | glucose:ti,ab,kw OR fdg:ti,ab,kw OR fluorodeoxyglucosef18:ti,ab,kw OR 18fluorodeoxyglucose:ti,ab,kw OR deoxyfluoroglucose:ti,ab,kw OR fludeoxyglucose:ti,ab,kw OR deoxyglucose:ti,ab,kw |
| #2 | fluorodeoxyglucose:ti,ab,kw |
| #1 | 'fluorodeoxyglucose f 18'/de |

**Scopus**

(TITLE-ABS-KEY ( fluorodeoxyglucose OR 18fluorodeoxyglucose OR fluorodeoxyglucosef18) OR TITLE-ABS-KEY ( fluoro W/2 deoxy W/2 glucose ) OR TITLE-ABS-KEY ( fdg OR deoxyfluoroglucose OR fludeoxyglucose OR deoxyglucose OR glucotrace OR glucose OR steripet ) OR CHEM ( 0z5b2cjx4d OR 63503-12-8 OR fluorodeoxyglucose )) AND (TITLE-ABS-KEY (Positron W/2 Emission W/2 Tomography) OR TITLE-ABS-KEY (PET)) AND ((TITLE-ABS-KEY(Amyotrophic W/2 Lateral W/2 Sclerosis) OR TITLE-ABS-KEY ("lou gehrig* disease" OR "guam disease")) OR (TITLE-ABS-KEY ("Parkinson* Disease" OR "paralysis agitans" OR "primary parkinsonism")) OR (TITLE-ABS-KEY((multiple or disseminated) W/2 sclerosis)) OR (TITLE-ABS-KEY("Alzheimer* Disease" OR "primary senile") OR TITLE-ABS-KEY(dementia W/2 (presenile or senile or alzheimer* or degenerative))))

**APA PsycInfo**

(DE "glucose metabolism" OR MA ( ("Fluorodeoxyglucose F18" OR glucose) ) OR TI ( fluorodeoxyglucose OR 18fluorodeoxyglucose OR fluorodeoxyglucosef18 OR (fluoro N2 deoxy N2 glucose) OR fdg OR deoxyfluoroglucose OR fludeoxyglucose OR deoxyglucose OR glucotrace OR glucose OR steripet ) OR AB ( fluorodeoxyglucose OR 18fluorodeoxyglucose OR fluorodeoxyglucosef18 OR (fluoro N2 deoxy N2 glucose) OR fdg OR deoxyfluoroglucose OR fludeoxyglucose OR deoxyglucose OR glucotrace OR glucose OR steripet ) OR KW ( fluorodeoxyglucose OR 18fluorodeoxyglucose OR fluorodeoxyglucosef18 OR (fluoro N2 deoxy N2 glucose) OR fdg OR deoxyfluoroglucose OR fludeoxyglucose OR deoxyglucose OR glucotrace OR glucose OR steripet )) AND (DE "Positron Emission Tomography" OR MA "Positron Emission Tomography" OR TI ( (Positron N2 Emission N2 Tomography) OR PET ) OR AB ( (Positron N2 Emission N2 Tomography) OR PET ) OR KW ( (Positron N2 Emission N2 Tomography) OR PET )) AND ( (DE "Amyotrophic lateral sclerosis" OR MA "Amyotrophic lateral sclerosis" OR TI ( (Amyotrophic N2 Lateral N2 Sclerosis) OR "lou gehrig* disease" OR "guam disease" ) OR AB ( (Amyotrophic N2 Lateral N2 Sclerosis) OR "lou gehrig* disease" OR "guam disease" ) OR KW ( (Amyotrophic N2 Lateral N2 Sclerosis) OR "lou gehrig* disease" OR "guam disease" )) OR (DE "Parkinson's Disease" OR MA "Parkinson Disease" OR TI ( "parkinson* disease" OR "paralysis agitans" OR "primary parkinsonism" ) OR AB ( "parkinson* disease" OR "paralysis agitans" OR "primary parkinsonism" ) OR KW ( "parkinson* disease" OR "paralysis agitans" OR "primary parkinsonism" )) OR (DE "Multiple sclerosis" OR MA "Multiple sclerosis" OR TI ( ((multiple or disseminated) N2 sclerosis) ) OR AB ( ((multiple or disseminated) N2 sclerosis) ) OR KW ( ((multiple or disseminated) N2 sclerosis) )) OR (DE "Alzheimer's Disease" OR MA "Alzheimer Disease" OR TI ( "Alzheimer* Disease" OR "primary senile" OR (dementia N2 (presenile or senile or alzheimer* or degenerative)) ) OR AB ( "Alzheimer* Disease" OR "primary senile" OR (dementia N2 (presenile or senile or alzheimer* or degenerative)) ) OR KW ( "Alzheimer* Disease" OR

"primary senile" OR (dementia N2 (presenile or senile or alzheimer* or degenerative)) ))
)

**Cochrane CENTRAL**

| ID | Search |
|---|---|
| #1 | [mh ^"Fluorodeoxyglucose F18"] |
| #2 | [mh ^glucose] |
| #3 | (fluorodeoxyglucose OR 18fluorodeoxyglucose OR fluorodeoxyglucosef18 OR (fluoro NEAR/2 deoxy NEAR/2 glucose) OR fdg OR deoxyfluoroglucose OR fludeoxyglucose OR deoxyglucose OR glucotrace OR glucose OR steripet):ti,ab,kw |
| #4 | {or #1-#3} |
| #5 | [mh ^"positron emission tomography"] |
| #6 | ((Positron NEAR/2 Emission NEAR/2 Tomography) OR PET):ti,ab,kw |
| #7 | {or #5-#6} |
| #8 | #4 AND #7 |
| #9 | [mh ^"Amyotrophic lateral sclerosis"] |
| #10 | ((Amyotrophic NEAR/2 Lateral NEAR/2 Sclerosis) OR "lou gehrig disease" OR "lou gehrig's disease" OR "guam disease"):ti,ab,kw |
| #11 | {or #9-#10} |
| #12 | #8 AND #11 |
| #13 | [mh ^"Parkinson Disease"] |
| #14 | ("parkinson disease" OR " parkinson's disease" OR "paralysis agitans" OR "primary parkinsonism"):ti,ab,kw |
| #15 | {or #13-#14} |
| #16 | #8 AND #15 |
| #17 | [mh ^"Multiple sclerosis"] |
| #18 | ((multiple or disseminated) NEAR/2 sclerosis):ti,ab,kw |
| #19 | {or #17-#18} |
| #20 | #8 AND #19 |
| #21 | [mh ^"Alzheimer Disease"] |
| #22 | ("Alzheimer Disease" OR " Alzheimer's Disease" OR "primary senile" OR (dementia NEAR/2 (presenile or senile or alzheimer* or degenerative)) ):ti,ab,kw |
| #23 | {or #21-#22} |
| #24 | #8 AND #23 |
| #25 | #12 OR #16 OR #20 OR #24 |
| #26 | Trials only |

**Supplemental Table 1. Reasons for full text exclusion**

| Exclusion Criteria | Full Text Excluded |
|---|---|
| Not late-onset neurodegeneration | 52 |
| Does not include a disease-free control group of individuals | 415 |
| Does not include 18F FDG-PET for quanitfying cerebral glucose metabolism | 13 |
| Data not analyzed in a voxelwise analysis | 529 |
| Data not data reported in a table with stereotactic coordinates, peak voxels, and a statistical outcome | 456 |
| Relevant disease group only includes mixed pathology (e.g. an "AD" group that includes both MCI and AD) | 35 |
| Outcomes only reported for region-of-interest analyses | 119 |
| Study group has early-onset neurodegenerative disease (e.g., familial, dominantly inherited, or diagnosis made prior to age 50) or atypical disease variants | 1 |
| Control group is not disease-free (e.g., other disease subtype) | 5 |
| FDG-PET data used for visual rating or diagnosis but not analyzed. | 2 |
| Data only analyzed at the single subject level, not group level | 1 |
| Outcomes only reported as PCA or other spatial decomposition method | 22 |
| Outcomes only reported as classifier (e.g., AD vs NC) | 6 |
| Review article or systematic review | 6 |
| Conference abstract or paper meeting a criterion for exclusion | 48 |
| Commentary without original results | 6 |
| Report contains insufficient information to assess methodological quality | 2 |
| Full report could not be retrieved | 95 |
| Clinical trial registration without data | 89 |

| Supplemental Table 2. Study level summary data | | | | | | |
|---|---|---|---|---|---|---|
| **Reference** | **Control sample size** | **$A^2$nd sample size** | **Mean control age** | **Mean A2nd age** | **No. of hypometabolic foci** | **No. of hypermetabolic foci** |
| **Studies reporting Alzheimer's disease outcomes** | | | | | | |
| Ibáñez et. al., 1998[105] | 19 | 15 | 69.7 | 68.9 | 10 | 0 |
| Ishii et. al., 2001[108] | 10 | 10 | 62 | 63 | 3 | 1 |
| Alexander et. al., 2002[54] | 34 | 14 | 64 | 65 | 30 | 0 |
| Jagust et. al., 2002[110] | 9 | 24 | 68.6 | 74.1 | 1 | 0 |
| Sakamoto et. al., 2002[153] | 17 | 20 | 73.9 | 75 | 2 | 0 |
| Volkow et. al., 2002[165] | 35 | 35 | 71.46 | 71.34 | 4 | 7 |
| Haier et. al., 2003[35] | 12 | 10 | Unreported | 76 | 15 | 11 |
| Eustache et. al., 2004[92] | 13 | 17 | 63.8 | 72.8 | 12 | 0 |
| Mosconi et. al., 2004[143] | 35 | 87 | 69.3 | 72 | 18 | 0 |
| Mosconi et. al., 2004[141] | 35 | 86† | 69.3 | 73.67 | 25 | 0 |
| Ouchi et. al., 2004[145] | 6 | 10 | 56.8 | 55.1 | 2 | 0 |
| Drzezga et. al., 2005[89] | 16 | 83† | 65 | 66.2 | 14 | 0 |
| Ishii et. al., 2005[107] | 30 | 30 | 66.8 | 66.8 | 2 | 0 |
| Kalpouzos et. al., 2005[115] | 15 | 26 | 62.4 | 75.2 | 5 | 0 |
| Kim et. al., 2005[119] | 13 | 46 | 71.5 | 72.8 | 1 | 0 |
| Sakamoto et. al., 2005[152] | 20 | 20 | 65.9 | 64.8 | 2 | 0 |
| Zahn et. al., 2005[174] | 10 | 10 | 65.8 | 66.5 | 11 | 0 |
| Kawachi et. al., 2006[118] | 30 | 32 | 66.6 | 67 | 6 | 0 |
| Teipel et. al., 2006[158] | 10 | 30 | 62 | 72.2 | 9 | 0 |
| Ziolko et. al., 2006[177] | 11 | 10 | 74 | 69 | 6 | 0 |
| Choo et. al., 2007[80] | 25 | 116† | 70 | 68.4 | 30 | 0 |
| Hunt et. al., 2007[104] | 14 | 44 | 65.5 | 69.3 | 17 | 0 |
| Ishii et. al., 2007[106] | 20 | 20 | 72.9 | 74.1 | 2 | 0 |
| Matsunari et. al., 2007[139] | 40 | 27 | 66.5 | 68.6 | 9 | 0 |
| Rauchs et. al., 2007[150] | 20 | 13 | 63.2 | 76.7 | 8 | 0 |
| Del Sole et. al., 2008[87] | 7 | 14 | Unreported | 75 | 12 | 0 |
| Drzezga et. al., 2008[90] | 26 | 8 | 64.5 | 73.5 | 13 | 0 |

| Supplemental Table 2. Study level summary data | | | | | | |
|---|---|---|---|---|---|---|
| **Reference** | **Control sample size** | **$A^2$nd sample size** | **Mean control age** | **Mean A2nd age** | **No. of hypometabolic foci** | **No. of hypermetabolic foci** |
| Habeck et. al., 2008[103] | 20 | 20 | 62 | 62 | 16 | 13 |
| Kanda et. al., 2008[116] | 20 | 20 | 65.2 | 65 | 10 | 7 |
| Lee et. al., 2008[124] | 40 | 71 | 70.6 | 68.6 | 8 | 0 |
| Mosconi et. al., 2008[142] | 47 | 6 | 65 | 72 | 6 | 0 |
| Langbaum et. al., 2009[121] | 82 | 74 | 68.4 | 71.1 | 13 | 0 |
| Yakushev et. al., 2009[169] | 15 | 36† | 66.2 | 69.7 | 7 | 0 |
| Chen et. al., 2010[72] | 79 | 69 | 76 | 75.3 | 13 | 0 |
| Laxton et. al., 2010[122] | 6 | 6 | 68.5 | 60.7 | 8 | 0 |
| Pascual et. al., 2010[148] | 12 | 12 | 79.2 | 78.2 | 5 | 0 |
| Shin et. al., 2010[154] | 10 | 10 | 70 | 73 | 16 | 0 |
| Teune et. al., 2010[36] | 18 | 15 | 56 | 65 | 9 | 12 |
| Yuan et. al., 2010[173] | 52* | 39† | 67.58 | 68.5 | 33 | 0 |
| Chen et. al., 2011[73] | 19 | 15 | 69.2 | 72 | 13 | 0 |
| Yokokura et. al., 2011[171] | 11 | 11 | Unreported | 70.6 | 3 | 0 |
| Pascual et. al., 2012[149] | 6 | 6 | 78.2 | 78 | 5 | 0 |
| Toussaint et. al., 2012[162] | 40 | 40 | 75.5 | 75.1 | 7 | 0 |
| Frisch et. al., 2013[97] | 13 | 19 | 53.92 | 60.89 | 2 | 0 |
| Küntzelmann et. al., 2013[120] | 10 | 15 | 58.8 | 69.8 | 1 | 6 |
| Castellano et. al., 2014[70] | 29 | 10 | 72 | 76 | 7 | 0 |
| Fu et. al., 2014[98] | 14 | 14 | 67.4 | 68.1 | 4 | 0 |
| Buchholz et. al., 2015[58] | 37 | 20 | 70 | 64.7 | 1 | 0 |
| Fan et. al., 2015[94] | 16 | 8 | 65 | 66.4 | 16 | 0 |
| Fan et. al., 2015[93] | 8 | 10 | 64.9 | 66.3 | 15 | 0 |
| Jedidi et. al., 2015[111] | 24 | 26 | 71.85 | 77.73 | 4 | 0 |
| Matias-Guiu et. al., 2015[138] | 9 | 33 | 60.5 | 74.2 | 7 | 0 |
| Verfaillie et. al., 2015[164] | 10 | 18 | 56 | 64 | 5 | 0 |

| Supplemental Table 2. Study level summary data | | | | | | |
|---|---|---|---|---|---|---|
| **Reference** | **Control sample size** | **A$^2$nd sample size** | **Mean control age** | **Mean A2nd age** | **No. of hypometabolic foci** | **No. of hypermetabolic foci** |
| Chiaravalloti et. al., 2016[78] | 58* | 84† | 67.9 | 68.25 | 7 | 0 |
| Liguori et. al., 2016[128] | 30 | 32 | 70.8 | 69.9 | 14 | 0 |
| Mattis et. al., 2016[29] | 20 | 20 | 76 | 76.6 | 11 | 7 |
| Aziz et. al., 2017[55] | 51* | 52† | 65 | 68.6 | 6 | 0 |
| Carapelle et. al., 2017[68] | 25 | 27 | 68.2 | 71.5 | 10 | 0 |
| Chiaravalloti et. al., 2017[79] | 13 | 87 | 71 | 70 | 7 | 0 |
| Fernández-Matarrubia et. al., 2017[95] | 24 | 29 | 67.4 | 75.3 | 5 | 0 |
| Chiaravalloti et. al., 2018[77] | 58 | 38 | 67 | 69 | 9 | 0 |
| Chiaravalloti et. al., 2018[76] | 20 | 131 | 67 | 70 | 10 | 0 |
| Fu et. al., 2018[99] | 14 | 14 | 67.4 | 68.1 | 21 | 0 |
| Weise et. al., 2018[168] | 40 | 51 | 76.1 | 75.2 | 15 | 0 |
| Chiaravalloti et. al., 2019[75] | 34 | 43 | 71 | 70 | 6 | 0 |
| Göttler et. al., 2019[101] | 22 | 32 | 63.6 | 65.9 | 4 | 0 |
| Li et. al., 2019[127] | 52* | 52† | 57.3 | 57.3 | 17 | 0 |
| Liguori et. al., 2019[129] | 35 | 55 | 67.89 | 69.18 | 15 | 0 |
| Luo et. al., 2019[133] | 27 | 63 | 73.5 | 74 | 2 | 1 |
| Meyer et. al., 2019[140] | 12 | 55† | Noted as "significantly younger" | 78.6 | 2 | 0 |
| Ceccarini et. al., 2020[71] | 30 | 8 | 63.9 | 70.88 | 9 | 0 |
| Gupta et. al., 2020[102] | 35 | 33 | 77.83 | 75.65 | 8 | 0 |
| Terada et. al., 2020[159] | 14 | 32† | 64.4 | 69.95 | 11 | 0 |
| Ye et. al., 2020[170] | 21 | 38 | 71.8 | 70.8 | 22 | 0 |
| Cappelletto et. al., 2021[67] | 50 | 60† | 62.32 | 72.85 | 24 | 0 |
| Chen et. al., 2021[74] | 24 | 23 | 54.67 | 58.74 | 3 | 0 |
| Carneiro et. al., 2022[69] | 24 | 27 | 71 | 74 | 2 | 1 |

| Supplemental Table 2. Study level summary data | | | | | | |
|---|---|---|---|---|---|---|
| **Reference** | **Control sample size** | **$A^2$nd sample size** | **Mean control age** | **Mean A2nd age** | **No. of hypometabolic foci** | **No. of hypermetabolic foci** |
| Cabrera-Martín et. al., 2023[60] | 60 | 180 | 71.03 | 72.84 | 26 | 0 |
| Dang et. al., 2023[84] | 38 | 41 | 64.32 | 72.59 | 11 | 0 |
| Liu et. al., 2023[130] | 33 | 50 | 55.82 | 58.86 | 4 | 0 |
| Lv et. al., 2023[134] | 26 | 35 | 48.46 | 58.23 | 5 | 0 |
| Frings et. al., 2024[96] | 13 | 26 | 68.3 | 62.6 | 8 | 0 |
| Ishii et. al., 2024[109] | 63 | 49 | 75.2 | 68.1 | 4 | 0 |
| Terstege et. al., 2025[160] | 29* | 58† | 75.43 | 75.47 | 6 | 0 |
| **Studies reporting Parkinson's disease outcomes** | | | | | | |
| Juh et. al., 2004[113] | 22 | 8 | 67.8 | 67.9 | 3 | 0 |
| Nagano-Saito et. al., 2004[144] | 13 | 19† | 66.2 | 66.8 | 10 | 8 |
| Juh et. al., 2005[114] | 22 | 8 | 67.8 | 67.9 | 6 | 0 |
| Del Olmo et. al., 2006[86] | 5 | 9 | 63.2 | 61 | 3 | 1 |
| Yong et. al., 2007[172] | 15 | 13 | 65.3 | 75.3 | 8 | 0 |
| Hosokai et. al., 2009[34] | 13 | 40† | 63 | 66.65 | 27 | 0 |
| Ma et. al., 2009[135] | 24 | 24 | 57 | 57.1 | 0 | 8 |
| Le Jeune et. al., 2010[123] | 13 | 20 | 53.23 | 57.9 | 3 | 5 |
| Teune et. al., 2010[36] | 18 | 20 | 56 | 63 | 14 | 9 |
| Berti et. al., 2012[56] | 21 | 26 | 62.4 | 65.3 | 8 | 8 |
| Garcia-Garcia et. al., 2012[100] | 20 | 115† | 67.9 | 71.73 | 53 | 0 |
| Edison et. al., 2013[91] | 8 | 6 | 65.9 | 68.2 | 9 | 0 |
| Fan et. al., 2015[93] | 8 | 11 | 64.9 | 68.4 | 15 | 0 |
| Chung et. al., 2016[82] | 15 | 24† | 65.7 | 64.75 | 14 | 0 |
| Zhang et. al., 2016[175] | 17 | 30† | 63.47 | 66.34 | 9 | 0 |
| Ko et. al., 2017[33] | 19 | 18 | 59.7 | 70.7 | 5 | 13 |
| Shin et. al., 2017[155] | 9 | 22† | 68 | 73.05 | 3 | 1 |
| Wang et. al., 2017[166] | 16 | 58 | 54.8 | 56.8 | 2 | 0 |
| Wang et. al., 2017[167] | 15 | 28† | 63.33 | 66.22 | 12 | 0 |

| Supplemental Table 2. Study level summary data | | | | | | |
|---|---|---|---|---|---|---|
| **Reference** | **Control sample size** | **A$^2$nd sample size** | **Mean control age** | **Mean A2nd age** | **No. of hypometabolic foci** | **No. of hypermetabolic foci** |
| Jin et. al., 2018[112] | 81 | 81 | 52.4 | 58.8 | 4 | 7 |
| Tan et. al., 2018[157] | 10 | 10 | 54.6 | 55.7 | 9 | 18 |
| Chu et. al., 2019[81] | 20 | 50 | 55.6 | 57.7 | 6 | 2 |
| Liguori et. al., 2019[129] | 35 | 28 | 67.89 | 65.6 | 3 | 0 |
| Kanno et. al., 2020[117] | 14 | 40† | 64 | 68 | 33 | 0 |
| Ruppert et. al., 2020[151] | 14 | 42 | 64.5 | 67.24 | 3 | 0 |
| Lu et. al., 2022[132] | 15 | 45† | 65.7 | 65.7 | 11 | 23 |
| Park et. al., 2022[147] | 7 | 12 | 67.9 | 63.5 | 2 | 0 |
| Steidel et. al., 2022[156] | 14 | 17 | 64.5 | 67.12 | 1 | 0 |
| Zhang et. al., 2022[176] | 16 | 18 | 54.8 | 58.4 | 11 | 15 |
| Biassoni et. al., 2023[57] | 42 | 44† | 69.6 | 72.47 | 22 | 0 |
| Li et. al., 2025[126] | 15 | 101† | 62.05 | 60.26 | 7 | 7 |
| Li et. al., 2025[125] | 40 | 80† | 66.12 | 63.38 | 10 | 17 |
| **Studies reporting ALS outcomes** | | | | | | |
| Cistaro et. al., 2012[32] | 22 | 32 | 62 | 63.3 | 8 | 11 |
| Cistaro et. al., 2014[83] | 40 | 30 | 62.2 | 58.2 | 65 | 3 |
| Pagani et. al., 2014[146] | 40 | 195 | 62 | 63.2 | 11 | 7 |
| Van Laere et. al., 2014[163] | 20 | 70 | 62.4 | 62.1 | 1 | 1 |
| Matías-Guiu et. al., 2016[138] | 24 | 18 | 60.5 | 57.89 | 8 | 4 |
| Buhour et. al., 2017[59] | 37 | 37 | 59 | 61.83 | 5 | 3 |
| Marini et. al., 2018[136] | 44 | 44 | 64 | 64 | 6 | 5 |
| Diehl-Schmid et. al., 2019[88] | 23 | 22 | 64.78 | 63.25 | 2 | 0 |
| Canosa et. al., 2020[63] | 40 | 131† | Unreported | 64.85 | 13 | 0 |
| Calvo et. al., 2022[61] | 40 | 79† | Unreported‡ | Unreported‡ | 15 | 0 |
| Canosa et. al., 2022[62] | 40 | 46 | Unreported | 65.4 | 32 | 0 |
| Canosa et. al., 2023[65] | 40 | 334† | 65.5 | 64 | 26 | 0 |
| De Vocht et. al., 2023[85] | 20 | 69† | 62.4 | 61.5 | 9 | 2 |

| **Supplemental Table 2. Study level summary data** | | | | | | |
|---|---|---|---|---|---|---|
| **Reference** | **Control sample size** | **$A^2$nd sample size** | **Mean control age** | **Mean A2nd age** | **No. of hypometabolic foci** | **No. of hypermetabolic foci** |
| Liu et. al., 2023[131] | 128 | 93 | 55.24 | 56.31 | 6 | 12 |
| Tondo et. al., 2024[161] | 125 | 9 | 65.78 | 62 | 7 | 0 |
| Canosa et. al., 2025[64] | 40 | 48 | 66.5 | 65.5 | 16 | 0 |
| Canosa et. al., 2025[66] | 168* | 260† | 62.5 | 66.95 | 19 | 5 |

*Total control sample size aggregated over multiple control groups.
† Total A2nd sample size aggregated over multiple groups.
‡Participants in FDG-PET analyses drawn from random sample within larger cohort meeting inclusion criteria.

| Supplementary Table 3. Spatial location, anatomical labeling and sensitivity metrics for ALE-identified clusters | | | | | | | | | | | | | | | | |
|---|---|---|---|---|---|---|---|---|---|---|---|---|---|---|---|---|
| **Analysis** | **Cluster #** | **Volume (mm³)** | **Cluster Z-value** | **COM X** | **COM Y** | **COM Z** | **Anatomical Label** | **Jackknife Mean** | **Jackknife Max** | **Supporting Studies (%)** | **Total Foci** | **FSN Median [2.5th –97.5th]** | **Censored Drawers** | **Stability Mean** | **Stability ≥0.5 (%)** | **Stability ≥0.8 (%)** |
| **AD Hypermetabolism (k=10)** | | | | | | | | | | | | | | | | |
| | 1 | 1760 | 2.83 | −50 | −14 | 28 | L Postcentral / Primary somatosensory cortex (BA3a) | 0.10 | 0.29 | 5 (50%) | 7 | 17.5 [5.0–50.0] | 2 | 0.38 | 34 | 0 |
| | 2 | 880 | 2.54 | 30 | −18 | −4 | R Putamen / Optic radiation | 0.10 | 0.40 | 3 (30%) | 3 | 17.1 [9.0–42.6] | 0 | 0.29 | 0 | 0 |
| | 3 | 776 | 1.95 | −20 | −18 | −4 | L Thalamus / Corticospinal tract | 0.10 | 0.35 | 3 (30%) | 3 | 3.4 [1.2–8.1] | 0 | 0.29 | 0 | 0 |
| **AD Hypometabolism (k=104)** | | | | | | | | | | | | | | | | |
| | 1 | 20736 | 3.72 | 0 | −52 | 34 | L Precuneus / Superior parietal lobule (7M) | 0.009 | 0.04 | 75 (72%) | 128 | 520.0 [520.0–520.0]† | 10 | 0.77 | 80 | 64 |
| | 2 | 14408 | 3.72 | −46 | −62 | 38 | L Angular / Inferior parietal lobule (PGa) | 0.009 | 0.04 | 58 (56%) | 79 | 520.0 [520.0–520.0]† | 10 | 0.64 | 66 | 43 |
| | 3 | 13776 | 3.72 | 50 | −58 | 38 | R Angular / Inferior parietal lobule (PGa) | 0.009 | 0.04 | 61 (59%) | 82 | 520.0 [520.0–520.0]† | 10 | 0.74 | 76 | 57 |
| | 4 | 6936 | 3.72 | −60 | −44 | −14 | L Temporal inf | 0.009 | 0.05 | 32 (31%) | 37 | 520.0 [520.0–520.0]† | 10 | 0.64 | 67 | 44 |
| | 5 | 4096 | 3.72 | 64 | −38 | −14 | R Temporal inf | 0.009 | 0.11 | 19 (18%) | 23 | 496.5 [436.8–520.0] | 6 | 0.65 | 71 | 41 |
| | 6 | 1752 | 2.82 | 40 | 24 | 44 | R Frontal mid | 0.010 | 0.13 | 10 (10%) | 11 | 46.1 [13.2–136.1] | 0 | 0.27 | 11 | 0 |
| | 7 | 1096 | 2.70 | −12 | 8 | 10 | L Caudate | 0.010 | 0.14 | 8 (8%) | 8 | 57.3 [40.1–85.9] | 0 | 0.38 | 26 | 0 |
| | 8 | 680 | 2.16 | −28 | 48 | 36 | L Frontal mid | 0.010 | 0.19 | 6 (6%) | 6 | 61.5 [21.5–129.6] | 0 | 0.17 | 0 | 0 |

| Supplementary Table 3. Spatial location, anatomical labeling and sensitivity metrics for ALE-identified clusters | | | | | | | | | | | | | | | | |
|---|---|---|---|---|---|---|---|---|---|---|---|---|---|---|---|---|
| **Analysis** | **Cluster #** | **Volume ($mm^3$)** | **Cluster Z-value** | **COM X** | **COM Y** | **COM Z** | **Anatomical Label** | **Jackknife Mean** | **Jackknife Max** | **Supporting Studies (%)** | **Total Foci** | **FSN Median [2.5th –97.5th]** | **Censored Drawers** | **Stability Mean** | **Stability ≥0.5 (%)** | **Stability ≥0.8 (%)** |
| **ALS Hypermetabolism (k=12)** | | | | | | | | | | | | | | | | |
| | 1 | 720 | 2.35 | −24 | −16 | −16 | L Hippocampus | 0.08 | 0.35 | 3 (25%) | 3 | 11.3 [4.2–40.5] | 0 | 0.38 | 0 | 0 |
| | 2 | 584 | 1.72 | 8 | −20 | −22 | Brain-stem / Corticospinal tract | 0.08 | 0.57 | 2 (17%) | 3 | 4.2 [0.0–29.0] | 0 | 0.55 | 100 | 0 |
| **ALS Hypometabolism (k=22)** | | | | | | | | | | | | | | | | |
| | 1 | 2016 | 3.72 | −38 | 18 | 50 | L Frontal mid / Broca's area (BA44) | 0.05 | 0.21 | 8 (36%) | 10 | 58.3 [30.0–110.0] | 2 | 0.48 | 40 | 10 |
| | 2 | 1928 | 3.72 | −36 | −80 | −2 | L Occipital mid / Visual cortex (V5) | 0.05 | 0.21 | 6 (27%) | 10 | 74.2 [21.7–110.0] | 4 | 0.48 | 41 | 0 |
| | 3 | 1384 | 3.72 | −52 | 4 | 40 | L Precentral / Premotor cortex (BA6) | 0.05 | 0.17 | 7 (32%) | 7 | 77.7 [45.7–110.0] | 2 | 0.57 | 59 | 25 |
| | 4 | 1288 | 3.72 | 46 | 10 | 34 | R Frontal inf oper / Broca's area (BA44) | 0.05 | 0.18 | 6 (27%) | 7 | 80.5 [61.9–110.0] | 2 | 0.60 | 63 | 24 |
| | 5 | 1080 | 3.43 | 6 | −84 | 12 | R Calcarine / Visual cortex v1 (BA17) | 0.05 | 0.22 | 5 (23%) | 5 | 66.0 [30.8–108.0] | 1 | 0.45 | 53 | 0 |
| | 6 | 936 | 3.43 | −42 | 32 | 20 | L Frontal inf tri / Broca's area (BA45) | 0.05 | 0.24 | 5 (23%) | 5 | 69.3 [25.5–110.0] | 2 | 0.46 | 54 | 0 |
| | 7 | 808 | 3.16 | 44 | 4 | 52 | R Frontal mid / Premotor cortex (BA6) | 0.05 | 0.26 | 4 (18%) | 4 | 43.4 [13.7–109.3] | 1 | 0.37 | 0 | 0 |
| **PD Hypermetabolism (k=20)** | | | | | | | | | | | | | | | | |
| | 1 | 2296 | 3.72 | 22 | −16 | 0 | R Thalamus / Corticospinal tract | 0.05 | 0.17 | 9 (45%) | 11 | 19.7 [12.2–33.1] | 0 | 0.45 | 35 | 0 |

| Supplementary Table 3. Spatial location, anatomical labeling and sensitivity metrics for ALE-identified clusters | | | | | | | | | | | | | | | | |
|---|---|---|---|---|---|---|---|---|---|---|---|---|---|---|---|---|
| **Analysis** | **Cluster #** | **Volume (mm³)** | **Cluster Z-value** | **COM X** | **COM Y** | **COM Z** | **Anatomical Label** | **Jackknife Mean** | **Jackknife Max** | **Supporting Studies (%)** | **Total Foci** | **FSN Median [2.5th –97.5th]** | **Censored Drawers** | **Stability Mean** | **Stability ≥0.5 (%)** | **Stability ≥0.8 (%)** |
| | 2 | 1792 | 3.72 | 10 | −48 | −20 | R Cerebelum 4 | 0.05 | 0.18 | 7 (35%) | 8 | 84.7 [55.9–100.0] | 6 | 0.62 | 69 | 32 |
| | 3 | 1144 | 2.13 | −20 | −18 | 2 | L Thalamus / Corticospinal tract | 0.05 | 0.23 | 5 (25%) | 5 | 5.8 [3.0–13.3] | 0 | 0.40 | 12 | 0 |
| | 4 | 1112 | 2.97 | −10 | −30 | 66 | L Paracentral lobule / Corticospinal tract | 0.05 | 0.24 | 5 (25%) | 5 | 31.2 [13.2–81.8] | 0 | 0.48 | 47 | 0 |
| | 5 | 816 | 2.32 | 20 | 14 | −14 | R Rectus / Inferior occipito-frontal fascicle | 0.05 | 0.33 | 4 (20%) | 4 | 21.9 [6.2–69.4] | 0 | 0.41 | 0 | 0 |
| **PD Hypometabolism (k=45)** | | | | | | | | | | | | | | | | |
| | 1 | 7312 | 3.72 | −44 | −68 | 34 | L Angular / Inferior parietal lobule (PGp) | 0.02 | 0.10 | 21 (47%) | 33 | 223.6 [214.2–225.0] | 9 | 0.53 | 49 | 25 |
| | 2 | 2672 | 3.72 | 42 | −66 | 42 | R Angular / Inferior parietal lobule (PGp) | 0.02 | 0.15 | 9 (20%) | 12 | 105.9 [50.1–212.0] | 1 | 0.44 | 40 | 0 |
| | 3 | 1408 | 3.29 | 46 | 22 | −12 | R Frontal inf orb | 0.02 | 0.18 | 7 (16%) | 7 | 73.1 [40.2–145.3] | 0 | 0.48 | 51 | 0 |
| | 4 | 1360 | 3.43 | −14 | 12 | 4 | L Caudate | 0.02 | 0.16 | 7 (16%) | 7 | 179.2 [84.7–225.0] | 5 | 0.51 | 54 | 0 |
| | 5 | 1320 | 3.43 | 16 | −96 | 0 | R Calcarine / Visual cortex v1 (BA17) | 0.02 | 0.18 | 7 (16%) | 7 | 82.1 [47.5–117.9] | 0 | 0.48 | 49 | 0 |
| | 6 | 888 | 2.53 | 14 | 12 | 2 | R Caudate / Callosal body | 0.02 | 0.22 | 5 (11%) | 5 | 51.6 [15.5–198.2] | 1 | 0.31 | 0 | 0 |
| | 7 | 752 | 2.18 | −8 | 8 | 68 | L Supp motor area / Premotor cortex (BA6) | 0.02 | 0.24 | 4 (9%) | 4 | 23.3 [7.2–66.0] | 0 | 0.26 | 0 | 0 |

| Supplementary Table 3. Spatial location, anatomical labeling and sensitivity metrics for ALE-identified clusters | | | | | | | | | | | | | | | | |
|---|---|---|---|---|---|---|---|---|---|---|---|---|---|---|---|---|
| **Analysis** | **Cluster #** | **Volume (mm³)** | **Cluster Z-value** | **COM X** | **COM Y** | **COM Z** | **Anatomical Label** | **Jackknife Mean** | **Jackknife Max** | **Supporting Studies (%)** | **Total Foci** | **FSN Median [2.5th –97.5th]** | **Censored Drawers** | **Stability Mean** | **Stability ≥0.5 (%)** | **Stability ≥0.8 (%)** |
| **A2ND Hypermetabolism (k=42)** | | | | | | | | | | | | | | | | |
| | 1 | 4080 | 3.72 | 24 | −16 | −4 | R Thalamus / Corticospinal tract | 0.02 | 0.10 | 17 (40%) | 19 | 128.9 [77.0–210.0] | 2 | 0.46 | 43 | 10 |
| | 2 | 2872 | 3.72 | −22 | −16 | −6 | L Thalamus / Corticospinal tract | 0.02 | 0.10 | 14 (33%) | 14 | 150.4 [91.7–210.0] | 3 | 0.52 | 52 | 11 |
| | 3 | 1488 | 3.43 | 10 | −46 | −20 | R Cerebelum 4 | 0.02 | 0.17 | 7 (17%) | 8 | 81.2 [44.7–192.2] | 1 | 0.42 | 42 | 0 |
| | 4 | 1384 | 3.12 | 36 | −2 | −26 | R Hippocampus / Amygdala | 0.02 | 0.17 | 7 (17%) | 7 | 62.5 [30.4–148.6] | 0 | 0.41 | 34 | 0 |
| | 5 | 992 | 2.46 | −8 | −30 | 64 | L Paracentral lobule / Corticospinal tract | 0.02 | 0.23 | 5 (12%) | 5 | 22.4 [15.2–35.8] | 0 | 0.36 | 0 | 0 |
| | 6 | 976 | 2.45 | 18 | 16 | −16 | R Rectus / Inferior occipito-frontal fascicle | 0.02 | 0.21 | 5 (12%) | 5 | 23.7 [14.2–73.5] | 0 | 0.35 | 0 | 0 |
| **A2ND Hypometabolism (k=171)** | | | | | | | | | | | | | | | | |
| | 1 | 18800 | 3.72 | −46 | −62 | 36 | L Angular / Inferior parietal lobule (PGp) | 0.006 | 0.03 | 87 (51%) | 131 | 855.0 [855.0–855.0]† | 10 | 0.65 | 65 | 46 |
| | 2 | 17752 | 3.72 | 0 | −52 | 34 | L Precuneus / Superior parietal lobule (7M) | 0.006 | 0.04 | 79 (46%) | 127 | 855.0 [855.0–855.0]† | 10 | 0.71 | 74 | 58 |
| | 3 | 15216 | 3.72 | 50 | −60 | 38 | R Angular / Inferior parietal lobule (PGa) | 0.006 | 0.03 | 78 (46%) | 111 | 855.0 [855.0–855.0]† | 10 | 0.74 | 76 | 61 |
| | 4 | 8496 | 3.72 | −60 | −48 | −12 | L Temporal inf | 0.006 | 0.04 | 52 (30%) | 63 | 855.0 [855.0–855.0]† | 10 | 0.59 | 59 | 41 |
| | 5 | 4064 | 3.72 | 64 | −38 | −14 | R Temporal inf | 0.006 | 0.08 | 23 (13%) | 28 | 761.4 [639.5–855.0] | 4 | 0.64 | 66 | 43 |
| | 6 | 4056 | 3.72 | 38 | 18 | 46 | R Frontal mid / Broca's area (BA44) | 0.006 | 0.05 | 28 (16%) | 34 | 67.9 [26.1–206.8] | 0 | 0.35 | 23 | 9 |

**Supplementary Table 3. Spatial location, anatomical labeling and sensitivity metrics for ALE-identified clusters**

| Analysis | Cluster # | Volume (mm³) | Cluster Z-value | COM X | COM Y | COM Z | Anatomical Label | Jackknife Mean | Jackknife Max | Supporting Studies (%) | Total Foci | FSN Median [2.5th –97.5th] | Censored Drawers | Stability Mean | Stability ≥0.5 (%) | Stability ≥0.8 (%) |
|---|---|---|---|---|---|---|---|---|---|---|---|---|---|---|---|---|
| | 7 | 2168 | 3.06 | −36 | 24 | 46 | L Frontal mid / Broca's area (BA44) | 0.006 | 0.11 | 16 (9%) | 20 | 134.1 [25.8–332.1] | 0 | 0.19 | 0 | 0 |
| | 8 | 1608 | 3.72 | −12 | 10 | 6 | L Caudate | 0.006 | 0.08 | 14 (8%) | 14 | 447.4 [261.7–816.5] | 1 | 0.58 | 64 | 37 |
| | 9 | 976 | 2.00 | 14 | −98 | 0 | R Calcarine / Visual cortex v1 (BA17) | 0.006 | 0.12 | 9 (5%) | 9 | 73.0 [16.0–243.8] | 0 | 0.22 | 0 | 0 |

**Notes:**

† Fail-safe N estimates were right-censored in all 10 drawers, indicating the cluster remained significant at the maximum search limit. The true fail-safe N exceeds the reported value.

Anatomical labels were assigned using the AAL3, Harvard-Oxford, and Jülich cytoarchitectonic atlases. L = left; R = right. Volume is reported in mm³. Coordinates (X, Y, Z) are in MNI space and correspond to the center-of-mass within each cluster. Jackknife Mean and Max report the mean and maximum proportional contribution of any single experiment to the cluster statistic. Supporting Studies indicates the number (and percentage) of experiments contributing at least one focus within the cluster. FSN = fail-safe N; the median number of simulated null experiments tolerated before the cluster no longer survived correction, with 2.5th–97.5th percentile range across 10 independent null-study drawers. Censored Drawers = number of drawers (out of 10) in which the cluster remained significant at the maximum search limit. Stability Mean = mean proportion of resampled subsets (75% of experiments) in which each cluster voxel survived correction. Stability ≥0.5 and ≥0.8 = percentage of cluster voxels reaching the respective stability threshold.

**Abbreviations:** AD = Alzheimer's disease; ALS = amyotrophic lateral sclerosis; PD = Parkinson's disease; $A^2ND$ = combined neurodegenerative diseases; k = number of experiments; FSN = fail-safe N; L = left; R = right; sup = superior division; inf = inferior division; BA = Brodmann area; GM = grey matter; WM = white matter.

**Supplemental Table 4. Spatial location and anatomical labeling of statistically significant clusters from conjunction analyses**

| Analysis | Cluster # | Volume (mm3) | Cluster Z-value | COM X | COM Y | COM Z | Anatomical Labels |
|---|---|---|---|---|---|---|---|
| Hypometabolism: AD and PD | 1 | 4416 | 3.78 | -46 | -66 | 32 | L Angular / Inferior parietal lobule (PGp) |
| | 2 | 2040 | 3.67 | 42 | -70 | 42 | R Angular / Inferior parietal lobule (PGp) |
| | 3 | 464 | 3.8 | -12 | 10 | 6 | L Caudate |
| | 4 | 8 | 3.16 | -46 | -60 | 18 | L Temporal mid / Inferior parietal lobule (PGp) |
| Hypermetabolism: AD and PD | 1 | 384 | 3.47 | -22 | -16 | -4 | L Thalamus / Corticospinal tract |
| | 2 | 184 | 3.31 | 28 | -14 | -2 | R Putamen / Optic radiation |

**Notes:**

Anatomical labels were assigned using the AAL3, Harvard-Oxford, and Jülich cytoarchitectonic atlases. L = left; R = right. Volume is reported in mm³. Coordinates (X, Y, Z) are in MNI space and correspond to the center-of-mass within each cluster.

Abbreviations: AD = Alzheimer's disease; PD = Parkinson's disease; L = left; R = right

| **Supplemental Table 5. Spatial location and anatomical labeling of statistically significant clusters from subtraction analyses** | | | | | | | |
|---|---|---|---|---|---|---|---|
| Analysis | Cluster # | Volume (mm3) | Cluster Z-value | COM X | COM Y | COM Z | Anatomical Labels |
| AD more hypometabolic than PD | 1 | 28408 | 3.82 | -4 | -50 | 30 | L Precuneus / Callosal body |
| PD more hypometabolic than AD | 1 | 22072 | 3.68 | 2 | -84 | 4 | L Calcarine / Visual cortex v1 (BA17) |
| | 2 | 5104 | 3.74 | 46 | 16 | -20 | R Temporal pole sup |
| | 3 | 2400 | 3.59 | -8 | 60 | 6 | L Frontal sup medial |
| | 4 | 1912 | 3.43 | -2 | 14 | 60 | L Supp motor area / Premotor cortex (BA6) |
| | 5 | 1712 | 3.56 | -48 | 28 | -4 | L Frontal inf tri / Broca's area (BA45) |
| | 6 | 1232 | 3.53 | -6 | -54 | 0 | Vermis 4 / Visual cortex v2 (BA18) |
| | 7 | 552 | 3.66 | 12 | 16 | -10 | R Caudate / Callosal body |
| | 8 | 280 | 3.36 | -8 | 6 | 56 | L Supp motor area / Premotor cortex (BA6) |
| AD more hypometabolic than ALS | 1 | 19256 | 3.77 | -10 | -54 | 28 | L Precuneus / Callosal body |
| | 2 | 4432 | 3.7 | 42 | -56 | 34 | R Angular / Inferior parietal lobule (PGa) |
| | 3 | 1920 | 3.52 | 56 | -36 | -20 | R Temporal inf |
| ALS more hypometabolic than AD | 1 | 37864 | 3.78 | -8 | -88 | 8 | L Calcarine / Visual cortex v1 (BA17) |
| | 2 | 8216 | 3.76 | -56 | -4 | 36 | L Precentral / Premotor cortex (BA6) |
| | 3 | 4552 | 3.66 | 44 | -4 | 50 | R Precentral / Premotor cortex (BA6) |
| | 4 | 4496 | 3.47 | -4 | -24 | 70 | L Paracentral lobule / Premotor cortex (BA6) |
| | 5 | 2752 | 3.67 | -38 | 30 | 10 | L Frontal inf tri / Broca's area (BA45) |
| | 6 | 1040 | 3.75 | -38 | 20 | 48 | L Frontal mid / Broca's area (BA44) |
| | 7 | 752 | 3.61 | 44 | 8 | 34 | R Frontal inf oper / Broca's area (BA44) |
| | 8 | 432 | 3.36 | -42 | 34 | -4 | L Frontal inf orb / Broca's area (BA45) |
| | 9 | 368 | 3.43 | 44 | -26 | 60 | R Postcentral / Primary somatosensory cortex (BA1) |
| ALS more hypometabolic than PD | 1 | 7048 | 3.75 | -52 | -4 | 40 | L Precentral / Premotor cortex (BA6) |
| | 2 | 1336 | 3.57 | -22 | 32 | 46 | L Frontal mid |
| | 3 | 1136 | 3.66 | 44 | -2 | 34 | R Precentral / Corticospinal tract |
| | 4 | 448 | 3.4 | -44 | -76 | -6 | L Occipital inf / Visual cortex (V5) |
| PD more hypermetabolic than ALS | 1 | 248 | 3.47 | 0 | -20 | 56 | L Paracentral lobule / Primary motor cortex (BA4a) |

**Notes:**

Anatomical labels were assigned using the AAL3, Harvard-Oxford, and Jülich cytoarchitectonic atlases. L = left; R = right. Volume is reported in mm³. Coordinates (X, Y, Z) are in MNI space and correspond to the center-of-mass within each cluster.

Abbreviations: AD = Alzheimer's disease; ALS = amyotrophic lateral sclerosis; PD = Parkinson's disease; L = left; R = right; BA = Brodmann area; oper = operculum; Supp = Supplemental
